# Probing the diabetes and colorectal cancer relationship using gene – environment interaction analyses

**DOI:** 10.1101/2022.10.16.22280490

**Authors:** Niki Dimou, Andre E. Kim, Orlagh Flanagan, Neil Murphy, Virginia Diez-Obrero, Anna Shcherbina, Elom K Aglago, Emmanouil Bouras, Peter T Campbell, Graham Casey, Steven Gallinger, Stephen B Gruber, Mark A Jenkins, Yi Lin, Victor Moreno, Edward Ruiz-Narvaez, Mariana C Stern, Yu Tian, Kostas K Tsilidis, Volker Arndt, Elizabeth L Barry, James W Baurley, Sonja I Berndt, Stéphane Bézieau, Stephanie A Bien, D Timothy Bishop, Hermann Brenner, Arif Budiarto, Robert Carreras-Torres, Tjeng Wawan Cenggoro, Andrew T Chan, Jenny Chang-Claude, Stephen J Chanock, Xuechen Chen, David V Conti, Christopher H Dampier, Matthew Devall, David Drew, Jane C Figueiredo, Graham G Giles, Andrea Gsur, Tabitha A Harrison, Akihisa Hidaka, Michael Hoffmeister, Jeroen R Huyghe, Kristina Jordahl, Eric Kawaguchi, Temitope O Keku, Susanna C Larsson, Loic Le Marchand, Juan Pablo Lewinger, Li Li, Bharuno Mahesworo, John Morrison, Polly A Newcomb, Christina C Newton, Mireia Obon-Santacana, Jennifer Ose, Rish K Pai, Julie R Palmer, Nick Papadimitrou, Bens Pardamean, Anita R Peoples, Paul D P Pharoah, Elizabeth A Platz, John D Potter, Gad Rennert, Peter C Scacheri, Robert E Schoen, Yu-Ru Su, Catherine M Tangen, Stephen N Thibodeau, Duncan C Thomas, Cornelia Ulrich, Caroline Y Um, Franzel JB van Duijnhoven, Kala Visvanathan, Pavel Vodicka, Ludmila Vodickova, Emily White, Alicja Wolk, Michael O Woods, Conghui Qu, Anshul Kundaje, Li Hsu, W. James Gauderman, Marc J Gunter, Ulrike Peters

## Abstract

Diabetes is an established risk factor for colorectal cancer; however, the mechanisms underlying this relationship are not fully understood and the role of genetic variation is unclear. We used data from 3 genetic consortia (CCFR, CORECT, GECCO; 31,318 colorectal cancer cases/41,499 controls) and undertook genome-wide gene-environment interaction analyses with colorectal cancer risk, including interaction tests of genetics(G)xdiabetes and joint testing of Gxdiabetes, G-colorectal cancer association and/or G-diabetes correlation (2,3-degrees of freedom joint tests; d.f.). Based on the joint tests, variant rs3802177 in *SLC30A8* (p-value_3-d.f_.:5.46×10^−11^; regulates phosphorylation of the insulin receptor and phosphatidylinositol-3 kinase activity) and rs9526201 in *LRCH1* (p-value_2-d.f_.:7.84×10^−09^; regulates T cell migration and Natural Killer Cell cytotoxicity) were associated with colorectal cancer. These results suggest that variation in genes related to insulin signalling and immune function may modify the association of diabetes with colorectal cancer and provide novel insights into the biology underlying the diabetes and colorectal cancer relationship.

## Introduction

Colorectal cancer is the third most common cancer globally with an estimated number of 1.9 million new cases in 2020 (1). The etiology of colorectal cancer involves a complex interplay between genetic and environmental determinants. Currently, around 140 genetic variants have been identified by genome-wide association studies (GWAS) explaining ∼12% of the variability in colorectal cancer risk (2, 3). However, limited research has been conducted to understand the interaction between genetic and environmental/lifestyle risk factors on the risk of colorectal cancer. Understanding how genetic variation may modify the association of environmental and lifestyle exposures with colorectal cancer risk may potentially uncover novel biological pathways underlying disease etiology and contribute to the development of prevention strategies.

Type 2 diabetes (T2D), the most common form of diabetes, is an established risk factor for colorectal cancer (4). The biological mechanisms that underlie the association between T2D and colorectal cancer risk are not fully understood but likely entail exposure to hyperinsulinemia and insulin resistance as well as hyperglycemia, which often precede onset of T2D (5). However, it is possible that other, yet-to-be recognized, molecular pathways mediate the T2D-colorectal cancer relationship.

Gene-environment interaction (GxE) studies have been employed to investigate whether genetic variants modify the association of diet, lifestyle, and drugs with colorectal cancer (6). A previous GxE analysis of diabetes and risk of colorectal cancer was limited by small sample size and was focused on candidate genes (7). To provide further insights into the molecular pathways of diabetes with colorectal cancer risk, we undertook a large-scale genome-wide GxE analysis that tested for interactions between common and rare variants and diabetes in 31,318 colorectal cancer cases and 41,499 controls.

## Methods

### Study participants

For this gene-environment interaction analysis, we used data from 48 studies described elsewhere (2, 3, 8, 9) (Supplementary Table 1). Briefly, we combined genetic and epidemiologic data from studies participating in the Colon Cancer Family Registry (CCFR), the Genetics and Epidemiology of Colorectal Cancer Consortium (GECCO), and the Colorectal Cancer Transdisciplinary Study (CORECT) with individuals of European ancestry. For cohort studies, nested case-control sets were assembled. Controls were matched on factors such as age, sex, race, and enrolment date or trial group, when applicable. Colorectal adenocarcinoma cases were confirmed by medical records, pathology reports, or death-certificate information. All studies were approved by the relevant research ethics committee or institutional review board.

**Table 1.** Statistically significant results of the gene-environment interaction analyses for diabetes and single genetic variants for colorectal cancer risk.

| Variant | Chr | BP Position | Closest Gene | Annotation | Reference allele | Alternate allele | Alternate allele frequency | Method | P (D\|G)<sup>a</sup> | P (E\|G)<sup>b</sup> | P (GxE)<sup>c</sup> | P-value<sup>d</sup><br>(Covariate set 1)<sup>e</sup> | P-value<sup>d</sup><br>(Covariate set 2)<sup>f</sup> |
| --- | --- | --- | --- | --- | --- | --- | --- | --- | --- | --- | --- | --- | --- |
| Primary GxE testing |  |  |  |  |  |  |  |  |  |  |  |  |  |
| rs3802177 | 8 | 118185025 | *SLC30A8* | 3' UTR | G | A | 0.31 | 3-d.f. joint test | 4.05x10<sup>-01</sup> | 4.90x10<sup>-10</sup> | 7.49x10<sup>-04</sup> | 5.46x10<sup>-11</sup> | 1.01x10<sup>-10</sup> |
| Secondary GxE testing |  |  |  |  |  |  |  |  |  |  |  |  |  |
| rs9526201 | 13 | 47191972 | *LRCH1* | intron | G | A | 0.82 | 2-d.f. joint test | 1.87x10<sup>-04</sup> | NA | 1.33x10<sup>-06</sup> | 7.84x10<sup>-09</sup> | 8.82x10<sup>-09</sup> |

Chr=chromosome; BP Position=base pair position based on NCBI Build 37; d.f.=degrees of freedom; UTR= Untranslated region; NA=not applicable
<sup>a</sup> P-value corresponds to the association between genetic variants and colorectal cancer.
<sup>b</sup> P-value corresponds to the association between genetic variants and diabetes.
<sup>c</sup> P-value corresponds to the interaction between genetic variants and diabetes on risk of colorectal cancer.
<sup>d</sup> P-values correspond to each method used to test for interactions between genetic variants and diabetes.
<sup>e</sup> Covariate set 1: age at baseline, sex, study, genotyping platform, and the first three principal components (Colorectal cancer cases=31,318, Controls=41,499).
<sup>f</sup> Covariate set 2: covariate set 1 plus additional adjustment for body mass index.

Analyses were limited to individuals of European ancestry, based on self-reported race and clustering of principal components with 1000 Genomes EUR superpopulation. We further excluded individuals based on cryptic relatedness or duplicates (prioritizing cases and/or individuals genotyped on the better platform), genotyping/imputation errors, non-colorectal cancer outcomes, and age outliers. Additionally, individuals were excluded if they had missing diabetes status. The final pooled sample size was 31,318 colorectal cancer cases and 41,499 controls.

### Harmonization of epidemiologic data

Information on demographics and potential risk factors were collected by self-report using in-person interviews and/or structured questionnaires (10). Individuals with diabetes were defined using a binary self-reported diagnosis of the disease. Data were collected and centralized at the GECCO coordinating center (Fred Hutchinson Cancer Research Center). Briefly, data harmonization consisted of a multi-step procedure, where common data elements (CDEs) were defined *a priori*. Study questionnaires and data dictionaries were examined and, through an iterative process of communication with data contributors, elements were mapped to these CDEs. Definitions, permissible values, and standardized coding were implemented into a single database via SAS and T-SQL. The resulting data were checked for errors and outlying values within and between studies.

### Genotyping, quality assurance/quality control and imputation

Detailed information on genotyping, imputation, and quality control are presented elsewhere (2, 3). In brief, genotyped variants were excluded based on deviation from Hardy-Weinberg Equilibrium (p-value<1×10^−4^), low call rate (<95-98%), discrepancies between reported and genotypic sex, discordant calls between duplicates. Autosomal variants of all studies were imputed to the Haplotype Reference Consortium (HRC) r1.1 (2016) panel using the University of Michigan Imputation Server (11) and converted into a binary format for data management and analyses using R package BinaryDosage (12). Imputed variants were excluded if they had low imputation quality (*R*^2^< 0.8). After quality control, a total of over 7.2 million variants were used for the gene-environment interaction analysis for common variants and 25,216 gene sets for rare variants (i.e. with minor allele frequency below 1%).

## Statistical methods

### Association of diabetes with colorectal cancer risk

To evaluate the main effect of diabetes on colorectal cancer risk, each study was analyzed separately using logistic regression models. Study-specific results were combined using a random-effects meta-analysis (Hartung-Knapp) to obtain summary odds ratios (ORs) and 95% confidence intervals (CIs) across studies (13). We calculated the heterogeneity p-values using Cochran’s Q statistic (14) and funnel plots were used to identify studies with outlying ORs for potential exclusion and sensitivity analyses.

### GxE analyses for common variants

We performed genome-wide interaction scans using GxEScanR (15). As it has been shown that 2-step testing can substantially improve statistical power for identifying interaction, the primary interaction analyses were based on the traditional logistic regression and two-step method (EDGE approach) (16). In addition, our primary analyses also include testing the joint association of main genetic effect on colorectal cancer, G-E association, and GxE interaction by the 3-degrees of freedom (d.f.) test (17). In secondary analyses, we explored the joint association of main genetic effect and GxE interaction by the 2-d.f. test (18, 19) and interactions between aggregated rare variant sets (allele frequency <1%) at the gene level and diabetes status (20).

Logistic regression models test for interaction by including an interaction term between a particular genetic variant genotype and diabetes status and testing *H*_0_: *β* _*GXE*_ = 0. Two-step methods can improve statistical power by implementing a filtering step to prioritize variants for interaction testing, alleviating the multiple testing burden of genome-wide association studies (GWAS). We implemented a hybrid two-step method that prioritizes potential interaction loci by weighting GxE tests (step 2) based on the ranks of an independent test statistic (step 1). Step 1 tests include a joint test referred to as the EDGE statistic (16) of the marginal association of each variant with risk of colorectal cancer (21) and the association between each variant with diabetes in the combined case-control sample (22). Our approach modifies the original weighted hypothesis testing framework (23) by accounting for linkage disequilibrium in controlling for type I error (24) (details are provided in the Supplementary Methods).

The joint 2-d.f. test evaluate simultaneously the main genetic effect and the GxE interaction and has been shown to improve power to detect susceptibility loci under a wide range of circumstances by accounting for GxE interactions (18, 19). This joint testing framework can be extended to additionally include the association between genetic variant and diabetes in the combined case-control sample to form a 3-d.f. test (17).

Our primary inferences are based on the standard 1-d.f. GxE test, the 2-step EDGE approach, and the 3-d.f. joint test, with a family-wise error rate for each set to 0.05/3 to control for multiple testing. We note that this approach is conservative as these testing approaches are somewhat correlated. In secondary analyses, we used the 2-d.f. test and a p-value <5×10^−8^ to declare statistical significance, with the qualification that these findings were secondary. All tests were two-sided.

Imputed variant dosages were modelled as continuous variables (25). All analyses were adjusted for age at baseline, sex, study/genotyping platform, and the first three principal components to account for potential population structure. Statistically significant interactions were further adjusted for body mass index (BMI) because it is a potential confounder in the diabetes-colorectal cancer association (26).

For statistically significant findings, we estimated stratified ORs by modelling the association between diabetes and colorectal cancer risk stratified by genotype and association of the per-allele increase in genotype and colorectal cancer risk stratified by diabetes status. We assessed the extent of genomic inflation by quantile-quantile (Q-Q) plots and by calculating the genomic inflation factor (lambda). As lambda scales according to sample size, we also calculated lambda_1000_, which scales the genomic inflation factor to an equivalent study of 1000 cases and 1000 controls (27, 28).

To present 2-d.f., 3-d.f. test, and two-step-method results, we created additional plots after removing known GWAS colorectal cancer loci (and variants in close proximity ±2MB with correlation *r*^2^>0.2) (2) to ensure the overall significance is not driven merely by the main genetic effect on colorectal cancer.

Regional plots for all statistically significant findings were generated using LocusZoom v1.3 (29). Measures of linkage disequilibrium (LD) were estimated using our controls. Possible eQTL relationships were explored using the Genotype-Tissue Expression (GTEx V8) and the University of Barcelona and University of Virginia genotyping and RNA sequencing project (BarcUVa-Seq) datasets (30). The BarcUVa-Seq data has data on diabetes status of 410 participants which we used to test interactions between the genetic variants and diabetes on gene expression.

### Prediction of regulatory impact of candidate non-coding variants

We used ATAC-seq, DNASE-seq, H3K27ac histone ChIP-seq, and H3K4me1 histone ChIP-seq datasets of primary tissue from healthy colon and tumor primary tissue samples from Scacheri et al. (31), as well as from three colorectal cancer cell lines (SW480, HCT116, COLO205). These datasets were processed through ENCODE ATAC-seq/DNASE-seq (32) and histone ChIP-seq pipelines (33) to perform alignment and peak calling. Dataset sources are indicated in Supplementary Table 2. -log10(p-value) tracks were extracted from the MACS2 step of the pipeline for visualization in genome browsers. Irreproducible Discovery Rate (IDR) (34) peak calls for ATAC-seq and DNASE-seq datasets, as well as naive overlap peak calls for histone ChIP-seq datasets, were determined from the ENCODE pipelines. The pyGenomeTracks (35) software package was used to visualize chromatin accessibility across the functional datasets and to plot -log10(p-value) signal tracks. Peaks across samples from the same assay were concatenated across datasets, cropped to within 200 bp centered on the peak summit, and merged using bedtools (36) merge.

**Table 2.**
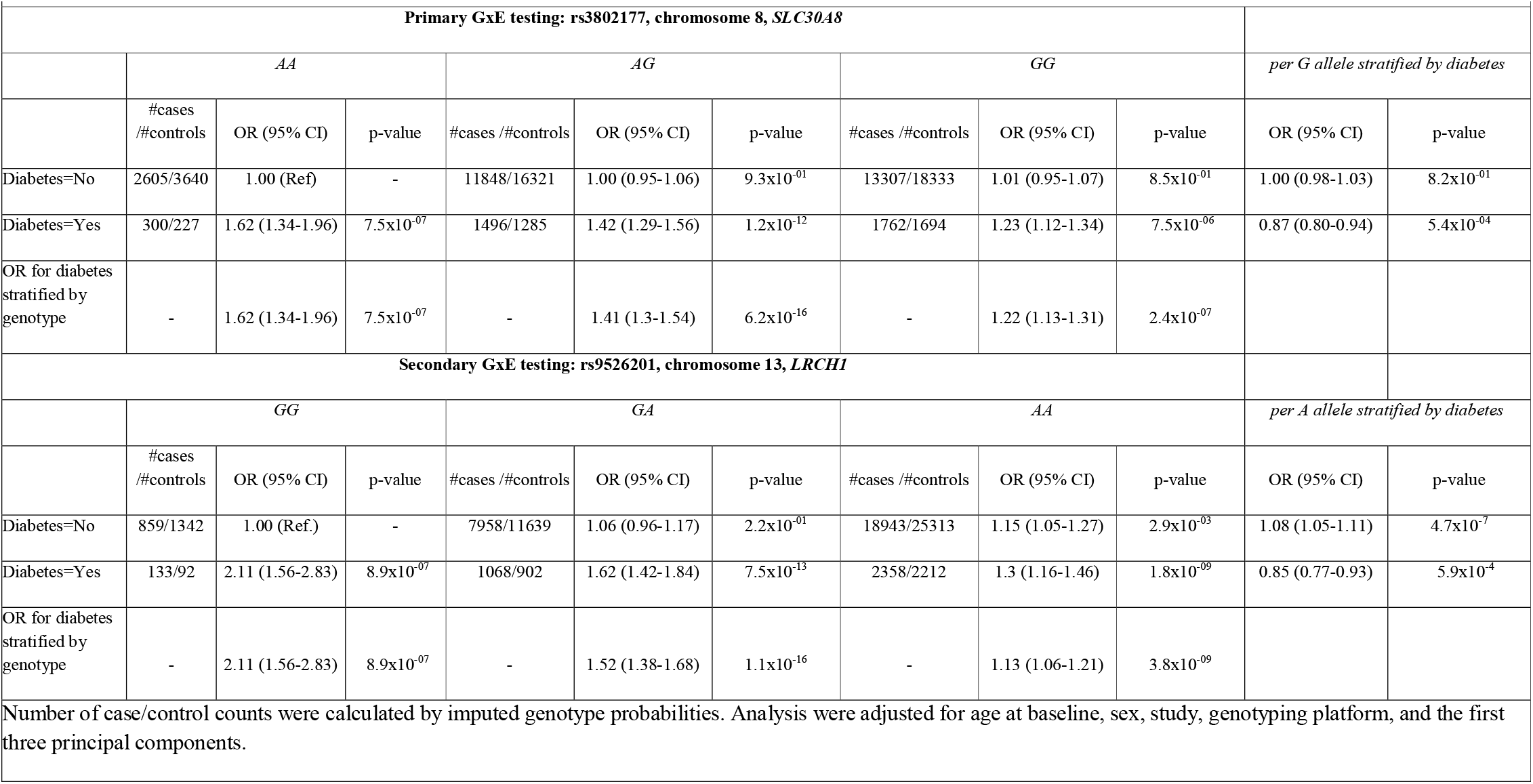
Stratified analysis for gene-diabetes interactions for colorectal cancer that were statistically significant. Odds ratios (OR) and 95% confidence interval (CI) are shown for association of diabetes with colorectal cancer risk stratified by genotype (rs3802177 and rs9526201) and for association of genotype with colorectal cancer risk stratified by diabetes.

Gapped k-mer support vector machine models (LS-GKM) (v0.1.0) with a center-weighted GKM kernel were trained to classify chromatin accessible regions against genomic background regions as a function of their underlying DNA sequences (37). Default parameters were utilized. Support vector machines (SVMs) were trained via 10-fold cross-validation, where groups of chromosomes were split into folds (Supplementary Table 3). Separate SVM models were trained on DNase-seq data from Supplementary Table 2 with samples pooled across assays as described above (31) (details are provided in the Supplementary Methods).

### GxE analyses for rare variants

As power for rare variant testing – and particularly GxE testing – tends to be low, we conducted GxE testing only for rare variants as a secondary analysis. We performed interaction tests of diabetes and aggregated rare variant sets at the gene and enhancer level (details are provided in the Supplementary Methods) using the Mixed effects Score Tests for interactions (MiSTi) method (20). This unified hierarchical regression framework combines the burdenxE as fixed effect and heterogeneous GxE effects as random effects. We considered a Fisher’s combination approach under MiSTi (fMiSTi) to discover GxE interactions (20), adjusting for age at baseline, sex, study, genotyping platform, and the first three principal components. Since 25,000 genes were tested and this was a secondary analysis, interactions with p-value<2×10^−6^ (*a*=0.05/25,000) were considered suggestively significant. The MiSTi R package was used for rare variants interaction analyses (20).

## Results

Overall, diabetes was associated with a significantly higher risk of colorectal cancer (OR: 1.36, 95% CI: 1.23-1.51, Supplementary Table 4), with similar results found in cohort and case-control studies. This association showed statistically significant between-study heterogeneity (Cochran’s Q p-value: <0.001; *I*^2^=47%, Supplementary Figure S1). However, there were no strong outlying studies (Supplementary Figure S2).

In our primary analysis we found that the association between diabetes and colorectal cancer risk is modified by variants on chromosome 8q24.11 within the *SLC30A8* gene based on the 3-d.f. joint test, with rs3802177 being the genetic variant showing the most significant effect (p-value: 5.46×10^−11^, Supplementary Figures S3A, S4A, Table 1). This result was robust in a sensitivity analysis accounting for BMI (Table 1). Although this variant was not directly associated with colorectal cancer (P-value: >0.05), we observed a strong association with diabetes (P-value: 4.90×10^−10^), and an interaction with diabetes for colorectal cancer risk (P-value: 7.49×10^−04^). When we stratified by genotype of rs3802177, we observed that the OR for diabetes vs. colorectal cancer among those carrying the *AA* genotype was the largest: 1.62, 95% CI: 1.34-1.96, P-value: 7.5×10^−07^, compared with OR: 1.41; 95% CI: 1.30-1.54; P-value: 6.2×10^−16^ among those carrying the *AG* genotype and, OR: 1.22; 95% CI: 1.13-1.31; P-value: 2.4×10^−07^ for those carrying the *GG* genotype. When stratifying by diabetes status, the risk of developing colorectal cancer per *G* allele was not statistically significant in those without diabetes (OR: 1.00; 95% CI: 0.98-1.03; P-value: 8.2×10^−01^) but was inverse among those with diabetes (OR: 0.87; 95% CI: 0.80-0.94; P-value: 5.4×10^−04^) (Table 2). We did not identify any statistically significant interactions using the traditional logistic regression or the 2-step approach. Genomic inflation for 1-d.f. GxE was minimal (lambda=1.008; lambda_1000_=1.000).

In our secondary 2-d.f. joint test, we identified that the association between diabetes and colorectal cancer risk is modified by a locus on chromosome 13q14.13 within the *LRCH1* gene, with genetic variant rs9526201 showing the most significant effect (p-value: 7.84×10^−09^, Supplementary Figures S3B, S4B, Table 1). This result was robust in a sensitivity analysis accounting for BMI (Table 1). As can be seen in Table 1, the p-value for the genetic variant-diabetes interaction was 1.33×10^−06^ and the association between genetic variants and colorectal cancer was 1.87×10^−04^, resulting in a combined significant 2-d.f. test statistic. When we stratified the association between diabetes and colorectal cancer by genotype of rs9526201, we observed a substantially stronger association among those carrying the *GG* genotype with an OR of 2.11 (95% CI: 1.56-2.83, P-value: 8.9×10^−07^), compared with an OR of 1.52 (95% CI: 1.38-1.68; P-value: 1.1×10^−16^) among those carrying the *GA* genotype and, an OR of 1.13 (95% CI: 1.06-1.21; P-value: 3.8×10^−09^) among those carrying the *AA* genotype. When stratifying by diabetes status, the risk of developing colorectal cancer increased per *A* allele in those without diabetes (OR: 1.08; 95% CI: 1.05-1.11; P-value: 4.7×10^−7^) but decreased in those with diabetes (OR: 0.85; 95% CI: 0.77-0.93; P-value: 5.9×10^−4^) (Table 2).

We did not identify any statistically significant GxE interactions when testing gene sets with rare variants.

We used two independent sources of eQTLs to evaluate the regulatory role of rs3802177 and rs9526201 variants on gene expression. Variant rs3802177 was not associated with gene expression in GTEx data; however, there was a suggestive eQTL in BarcUVa-Seq data that regulates expression of *AARD*, with the *G* allele associated with increased expression (β: 0.14, P-value: 4.7×10^−2^) (Supplementary Table S5). Also, variants in LD *R*^2^> 0.5 with rs3802177 were suggestive eQTLs in GTEx transverse colon data that are associated with the expression of *AARD* (Supplementary Table S6). For the BarcUVa data, we assessed the diabetes status of participants who provided this information (N: 49 individuals with diabetes; N: 361 without diabetes) and tested for interactions between the variant and diabetes on gene expression. There was no evidence of a statistically significant interaction (P-values> 0.05) of variant rs3802177 (or variants in LD *R*^2^> 0.5 with rs3802177) with diabetes in relation to *SLC30A8* gene expression in BarcUVa-Seq data (or any gene within 1Mb of rs3802177).

Variant rs9526201 is an eQTL in the GTEx V8 compendium that influences the expression of *LRCH1* in 8 non-colorectal tissues (Supplementary Table S7) and variants correlated with rs9526201 are suggestive eQTLs for *LRCH1* based on GTEx transverse colon tissue (Supplementary Table S6). Also, variant rs9526201 is a suggestive eQTL in normal colon tissue (from the BarcUVa-Seq data) that regulates expression of *RUBCNL*, with the *A* allele associated with increased expression (β: 0.17, p-value: 1.3×10^−2^) (Supplementary Table S5). We found a suggestive interaction (P-value: 0.02) of variant rs9534444 (LD *R*^2^: 0.52 with rs9526201) with diabetes in relation to *LRCH1* gene expression (Supplementary Figure S5).

Functional annotation analyses showed no evidence of enhancer activity for the variant rs3802177 or variants correlated with this variant (Supplementary Figure S6A). However, the variant rs9526201 in the *LRCH1* gene is associated with pronounced enhancer activity in colon tumor and cancer cell lines (Supplementary Figure S6B) and is in proximity with several variants that are located in open chromatin, suggesting enhancer activity in normal colon tissues, colorectal cancer cell lines, and several tissues (Supplementary Table S8).

We expanded our candidate set of variants to include variants in LD, in a 500 kb window around rs3802177 and rs9526201 variants (LD *R*^2^ >0.20) and used gkmSVM models to predict variant allelic effects on chromatin accessibility (Supplementary Figure S7). For rs3802177 and rs9526201, the models showed a weak difference in predicted chromatin accessibility between the reference and alternate alleles (Supplementary Table S9). We found a borderline difference in predicted chromatin accessibility between the alternate G allele and the reference C (ISM score= -1.148 in HCT116) for variant rs9534444 (LD *R*^2^: 0.52 with rs9526201; Supplementary Table S9).

GkmExplain analysis of rs3802177 and rs9526201 showed that there a was weak allelic effect in healthy tissue, tumor tissue, and cancer cell lines (Supplementary Figure S8). G to A variation in rs3802177 disrupts IRX3, leading to an increased probability of chromatin accessibility with the A allele whereas A to G allelic variation in rs9526201 completes a RUNX1 motif, leading to decreased probability of chromatin accessibility with the G allele (Supplementary Figure S8). For variant rs9534444 which is the highest-effect variant, in LD with rs9526201, results suggested that C to G variation disrupts motifs ZN341, MYF5, and PRGR.

## Discussion

In this large genome-wide GxE interaction analysis involving more than 30,000 colorectal cancer cases, we found that the association of diabetes status with colorectal cancer is modified by common genetic variants located within the *SLC30A8* and *LRCH1* genes. The mechanisms linking diabetes with colorectal cancer are not fully understood. Dysregulation of insulin and glucose metabolism are important candidate mechanisms and hyperinsulinemia itself has been causally linked to colorectal cancer development (38, 39); however, the precise mechanisms linking these phenomena are not clear. The findings of this analysis may provide biological insights into the established link between diabetes and colorectal cancer.

We found that the association of diabetes with colorectal cancer risk is modified by variants located in the *SLC30A8* gene. These genetic variants were not statistically significantly associated with gene expression in GTEx and only a weak eQTL has been observed in colorectal tissue for the *AARD* gene. Furthermore, the genetic variants were not located within predicted enhancer regions and we observed only weak evidence for allele-specific effects. Given the limited functional evidence, we focused on the closest gene, *SLC30A8*, which encodes a zinc transporter, ZnT8, that regulates zinc accumulation in the beta cells of the pancreas (40). Zinc is implicated in the phosphorylation of the insulin receptor beta-subunit and phosphatidylinositol 3-kinase (PI3K)/serine/threonine-specific protein kinase (Akt) signalling pathway (41, 42). Dysregulation of the PI3K/AKT pathway is associated with diabetes development (43) and with anti-apoptotic effects in colorectal cancer cells (44, 45). Our top hit, rs3802177 in the *SLC30A8* gene is in LD (*R*^2^ =1) with rs13266634, which was associated with diabetes risk in a previous GWAS (46) (as well as in our analysis) and has also been shown to modify insulin secretion (47). In summary, although we did not find strong functional genomic support for this highly significant association, the genetic variant is located within *SLC30A8* which is a strong candidate gene for modifying the diabetes-colorectal cancer association.

We also observed that the association of diabetes with colorectal cancer risk is modified by genetic variants located in *LRCH1*. eQTL as well as gene-expression analysis for the variantxdiabetes interaction suggests that *LRCH1* might represent the target gene regulating expression and transcription. The variants in LD with rs9526201 are located in enhancer peaks and we observed borderline significant allele-specific effects for variant rs9534444. For rs9534444, a C-to-G mutation disrupts the motif “TGGAAGAGCAGATGG”, which the TomTom software presents as a significant match to the know binding motifs of the ZN341, MYF5, PRGR transcription factors. The loss of function in response to the C-to-G mutation was observed in all 5 datasets profiled via SVM, with the strongest effects observed in the HCT116 cell line. LRCH1 is known to interact with DOCK8 to restrain the guanine-nucleotide exchange factor activity of DOCK8, resulting in the inhibition of Cdc42 activation and T cell migration (48). Cdc42 activation has been related to several malignancies, including colorectal cancer (49). Increased Cdc42 levels have been associated with colorectal cancer progression by promoting colorectal cancer cell migration and invasion (50) and regulating the putative tumor suppressor gene *ID4* (51). Low LRCH1 levels, which increase migration of CD4+ T cells, have also been found in patients with ulcerative colitis (52). Moreover, Cdc42 is implicated in Natural Killer (NK) cell cytotoxicity: Wiskott-Aldrich Syndrome protein which is the effector of Cdc42 is required for NK cell killing activity (53). Experimental evidence has shown that LRCH1 may regulate NK-92 cell cytotoxicity (54). Of further relevance to our finding, Cdc42 is implicated in insulin secretion and is linked to insulin resistance and diabetic nephropathy (55). One of the proposed mechanisms is via the Cdc42-p21-activated kinase1 (PAK1) signalling pathway essential for insulin secretion in human islets, as it was shown that individuals with diabetes were more likely to have an abnormal component of PAK1 (56). These data demonstrate a link between *LRCH1* and immune function via Cdc42 that is related to colorectal cancer and diabetes, which may explain the observed differential association. However, functional follow-up studies are needed to further explore this potential significant finding.

To our knowledge, there has been one previous study examining the interaction between T2D genetic variants and diabetes status in colorectal cancer risk, which included 1,798 colorectal cancer cases and 1,810 controls and focused on T2D-related variants (7). That study found a statistically significant interaction of T2D with an intronic variant rs4402960 located at the *IGF2BP2* gene (interaction P-value: 0.040) and a missense variant rs1801282 at the *PPARG* gene (interaction P-value: 0.036) The respective p-values for interaction for rs4402960 and rs1801282 with diabetes on colorectal cancer were not nominally significant in our GxE analysis providing limited support for those previously observed interactions. Additionally, we previously conducted an analysis among a large subset of our studies (26,017 cases and 20,692 controls) evaluating interactions between genetic predicted gene-expression levels and diabetes on colorectal cancer risk, and identified a statistically significant interaction between genetically predicted gene expression levels for *PTPN2* and diabetes (P-value: 2.31×10^−5^) (57). As the approach of this previous analysis was use of multiple common variants to predict gene expression, we would not expect to replicate those findings here.

Strengths of this study include the large sample size and state of the art statistical approaches, including 2-step (16) and joint tests (17-19), that improved statistical power by leveraging direct gene-diabetes and gene-colorectal cancer associations induced by Gxdiabetes effects on colorectal cancer risk. We applied strict corrections to account for multiple comparisons because of the number of the methods used. We acknowledge that our results were limited to Europeans and thus our findings cannot be readily generalized to other populations but require to follow up in those population groups. In addition, our novel findings need to explored in experimental models. Also, we could not account for diabetes history and treatment which may have an effect on colorectal cancer risk. For example, an inverse association between metformin use and colorectal cancer risk has been found in some studies, but not all, while a clinical trial conducted in Japan reported a protective effect of metformin on colorectal polyp development (58). Future studies may also focus on incorporating data on pre-diabetes states and those with hyperinsulinemia.

In summary, our results suggest that variation in genes related to immune function and regulation of the insulin receptor and PI3K activity may modify the association between diabetes and colorectal cancer risk. These results provide novel insights into the biology underlying diabetes and colorectal cancer relationship. Further experimental studies are warranted to understand the mechanisms by which these genes play a role in linking diabetes and colorectal cancer development.

## Data Availability

The data underlying this article will be deposited into a public repository and the accession codes will be available before publication.

## Author contributions

N. Dimou: Methodology, writing-original draft, writing–review and editing; A.E. Kim, V. Diez-Obrero, A. Shcherbina: Formal analysis, methodology, writing-original draft, writing-review and editing; O. Flanagan: writing–original draft, writing–review and editing; A. Kundaje, L. Hsu, W.J. Gauderman, M.J. Gunter, U. Peters: Conceptualization, supervision, investigation, methodology, writing–original draft, writing–review and editing; All authors contributed to the refinement and revision of the paper.

## Code availability

The code used to generate results of this article will be shared on reasonable request to the corresponding author

## Funding

### Genetics and Epidemiology of Colorectal Cancer Consortium (GECCO)

National Cancer Institute, National Institutes of Health, U.S. Department of Health and Human Services (U01 CA137088, R01 CA059045, U01 CA164930, R01 CA206279, R01 CA201407). Genotyping/Sequencing services were provided by the Center for Inherited Disease Research (CIDR) contract number HHSN268201700006I and HHSN268201200008I. This research was funded in part through the NIH/NCI Cancer Center Support Grant P30 CA015704. Scientific Computing Infrastructure at Fred Hutch funded by ORIP grant S10OD028685.

ASTERISK

### a Hospital Clinical Research Program (PHRC-BRD09/C) from the University Hospital Center of Nantes (CHU de Nantes) and supported by the Regional Council of Pays de la Loire, the Groupement des Entreprises Françaises dans la Lutte contre le Cancer (GEFLUC), the Association Anne de Bretagne Génétique and the Ligue Régionale Contre le Cancer (LRCC)

CLUE II funding was from the National Cancer Institute (U01 CA086308, Early Detection Research Network; P30 CA006973), National Institute on Aging (U01 AG018033), and the American Institute for Cancer Research. The content of this publication does not necessarily reflect the views or policies of the Department of Health and Human Services, nor does mention of trade names, commercial products, or organizations imply endorsement by the US government. Maryland Cancer Registry (MCR) cancer data was provided by the Maryland Cancer Registry, Center for Cancer Prevention and Control, Maryland Department of Health, with funding from the State of Maryland and the Maryland Cigarette Restitution Fund. The collection and availability of cancer registry data is also supported by the Cooperative Agreement NU58DP006333, funded by the Centers for Disease Control and Prevention. Its contents are solely the responsibility of the authors and do not necessarily represent the official views of the Centers for Disease Control and Prevention or the Department of Health and Human Services.

The Colon Cancer Family Registry (CCFR, www.coloncfr.org) is supported in part by funding from the National Cancer Institute (NCI), National Institutes of Health (NIH) (award U01 CA167551). Support for case ascertainment was provided in part from the Surveillance, Epidemiology, and End Results (SEER) Program and the following U.S. state cancer registries: AZ, CO, MN, NC, NH; and by the Victoria Cancer Registry (Australia) and Ontario Cancer Registry (Canada). The CCFR Set-1 (Illumina 1M/1M-Duo) and Set-2 (Illumina Omni1-Quad) scans were supported by NIH awards U01 CA122839 and R01 CA143237 (to GC). The CCFR Set-3 (Affymetrix Axiom CORECT Set array) was supported by NIH award U19 CA148107 and R01 CA81488 (to SBG). The CCFR Set-4 (Illumina OncoArray 600K SNP array) was supported by NIH award U19 CA148107 (to SBG) and by the Center for Inherited Disease Research (CIDR), which is funded by the NIH to the Johns Hopkins University, contract number HHSN268201200008I. Additional funding for the OFCCR/ARCTIC was through award GL201-043 from the Ontario Research Fund (to BWZ), award 112746 from the Canadian Institutes of Health Research (to TJH), through a Cancer Risk Evaluation (CaRE) Program grant from the Canadian Cancer Society (to SG), and through generous support from the Ontario Ministry of Research and Innovation. The SFCCR Illumina HumanCytoSNP array was supported in part through NCI/NIH awards U01/U24 CA074794 and R01 CA076366 (to PAN). The content of this manuscript does not necessarily reflect the views or policies of the NCI, NIH or any of the collaborating centers in the Colon Cancer Family Registry (CCFR), nor does mention of trade names, commercial products, or organizations imply endorsement by the US Government, any cancer registry, or the CCFR.

### COLON

The COLON study is sponsored by Wereld Kanker Onderzoek Fonds, including funds from grant 2014/1179 as part of the World Cancer Research Fund International Regular Grant Programme, by Alpe d’Huzes and the Dutch Cancer Society (UM 2012–5653, UW 2013-5927, UW2015-7946), and by TRANSCAN (JTC2012-MetaboCCC, JTC2013-FOCUS). The Nqplus study is sponsored by a ZonMW investment grant (98-10030); by PREVIEW, the project PREVention of diabetes through lifestyle intervention and population studies in Europe and around the World (PREVIEW) project which received funding from the European Union Seventh Framework Programme (FP7/2007–2013) under grant no. 312057; by funds from TI Food and Nutrition (cardiovascular health theme), a public–private partnership on precompetitive research in food and nutrition; and by FOODBALL, the Food Biomarker Alliance, a project from JPI Healthy Diet for a Healthy Life.

### Colorectal Cancer Transdisciplinary (CORECT) Study

The CORECT Study was supported by the National Cancer Institute, National Institutes of Health (NCI/NIH), U.S. Department of Health and Human Services (grant numbers U19 CA148107, R01 CA081488, P30 CA014089, R01 CA197350; P01 CA196569; R01 CA201407; R01 CA242218), National Institutes of Environmental Health Sciences, National Institutes of Health (grant number T32 ES013678) and a generous gift from Daniel and Maryann Fong.

### CORSA

The CORSA study was funded by Austrian Research Funding Agency (FFG) BRIDGE (grant 829675, to Andrea Gsur), the “Herzfelder’sche Familienstiftung” (grant to Andrea Gsur) and was supported by COST Action BM1206.

### CPS-II

The American Cancer Society funds the creation, maintenance, and updating of the Cancer Prevention Study-II (CPS-II) cohort. This study was conducted with Institutional Review Board approval.

### CRCGEN

Colorectal Cancer Genetics & Genomics, Spanish study was supported by Instituto de Salud Carlos III, co-funded by FEDER funds –a way to build Europe– (grants PI14-613 and PI09-1286), Agency for Management of University and Research Grants (AGAUR) of the Catalan Government (grant 2017SGR723), Junta de Castilla y León (grant LE22A10-2), the Spanish Association Against Cancer (AECC) Scientific Foundation grant GCTRA18022MORE and the Consortium for Biomedical Research in Epidemiology and Public Health (CIBERESP), action Genrisk. Sample collection of this work was supported by the Xarxa de Bancs de Tumors de Catalunya sponsored by Pla Director d’Oncología de Catalunya (XBTC), Plataforma Biobancos PT13/0010/0013 and ICOBIOBANC, sponsored by the Catalan Institute of Oncology. We thank CERCA Programme, Generalitat de Catalunya for institutional support.

### Czech Republic CCS

This work was supported by the Grant Agency of the Czech Republic (18-09709S, 20-03997S), by the Grant Agency of the Ministry of Health of the Czech Republic (grants AZV NV18/03/00199 and AZV NV19-09-00237), and Charles University grants Unce/Med/006 and Progress Q28/LF1.

### DACHS

This work was supported by the German Research Council (BR 1704/6-1, BR 1704/6-3, BR 1704/6-4, CH 117/1-1, HO 5117/2-1, HE 5998/2-1, KL 2354/3-1, RO 2270/8-1 and BR 1704/17-1), the Interdisciplinary Research Program of the National Center for Tumor Diseases (NCT), Germany, and the German Federal Ministry of Education and Research (01KH0404, 01ER0814, 01ER0815, 01ER1505A and 01ER1505B).

### EPIC

The coordination of EPIC is financially supported by International Agency for Research on Cancer (IARC) and also by the Department of Epidemiology and Biostatistics, School of Public Health, Imperial College London which has additional infrastructure support provided by the NIHR Imperial Biomedical Research Centre (BRC). The national cohorts are supported by: Danish Cancer Society (Denmark); Ligue Contre le Cancer, Institut Gustave Roussy, Mutuelle Générale de l’Education Nationale, Institut National de la Santé et de la Recherche Médicale (INSERM) (France); German Cancer Aid, German Cancer Research Center (DKFZ), German Institute of Human Nutrition Potsdam-Rehbruecke (DIfE), Federal Ministry of Education and Research (BMBF) (Germany); Associazione Italiana per la Ricerca sul Cancro-AIRC-Italy, Compagnia di SanPaolo and National Research Council (Italy); Dutch Ministry of Public Health, Welfare and Sports (VWS), Netherlands Cancer Registry (NKR), LK Research Funds, Dutch Prevention Funds, Dutch ZON (Zorg Onderzoek Nederland), World Cancer Research Fund (WCRF), Statistics Netherlands (The Netherlands); Health Research Fund (FIS) - Instituto de Salud Carlos III (ISCIII), Regional Governments of Andalucía, Asturias, Basque Country, Murcia and Navarra, and the Catalan Institute of Oncology - ICO (Spain); Swedish Cancer Society, Swedish Research Council and County Councils of Skåne and Västerbotten (Sweden); Cancer Research UK (14136 to EPIC-Norfolk; C8221/A29017 to EPIC-Oxford), Medical Research Council (1000143 to EPIC-Norfolk; MR/M012190/1 to EPIC-Oxford). (United Kingdom).

ESTHER/VERDI. This work was supported by grants from the Baden-Württemberg Ministry of Science, Research and Arts and the German Cancer Aid.

### Harvard cohorts

HPFS is supported by the National Institutes of Health (P01 CA055075, UM1 CA167552, U01 CA167552, R01 CA137178, R01 CA151993, and R35 CA197735), NHS by the National Institutes of Health (P01 CA087969, UM1 CA186107, R01 CA137178, R01 CA151993, and R35 CA197735), and PHS by the National Institutes of Health (R01 CA042182).

### Hawaii Adenoma Study

NCI grants R01 CA072520.

### Kentucky

This work was supported by the following grant support: Clinical Investigator Award from Damon Runyon Cancer Research Foundation (CI-8); NCI R01CA136726.

### LCCS

The Leeds Colorectal Cancer Study was funded by the Food Standards Agency and Cancer Research UK Programme Award (C588/A19167).

MCCS cohort recruitment was funded by VicHealth and Cancer Council Victoria. The MCCS was further supported by Australian NHMRC grants 509348, 209057, 251553 and 504711 and by infrastructure provided by Cancer Council Victoria. Cases and their vital status were ascertained through the Victorian Cancer Registry (VCR) and the Australian Institute of Health and Welfare (AIHW), including the National Death Index and the Australian Cancer Database.

### MEC

National Institutes of Health (R37 CA054281, P01 CA033619, and R01 CA063464).

### NCCCS I & II

We acknowledge funding support for this project from the National Institutes of Health, R01 CA066635 and P30 DK034987.

### NFCCR

This work was supported by an Interdisciplinary Health Research Team award from the Canadian Institutes of Health Research (CRT 43821); the National Institutes of Health, U.S. Department of Health and Human Serivces (U01 CA074783); and National Cancer Institute of Canada grants (18223 and 18226). The authors wish to acknowledge the contribution of Alexandre Belisle and the genotyping team of the McGill University and Génome Québec Innovation Centre, Montréal, Canada, for genotyping the Sequenom panel in the NFCCR samples. Funding was provided to Michael O. Woods by the Canadian Cancer Society Research Institute.

### PLCO

Intramural Research Program of the Division of Cancer Epidemiology and Genetics and supported by contracts from the Division of Cancer Prevention, National Cancer Institute, NIH, DHHS. Funding was provided by National Institutes of Health (NIH), Genes, Environment and Health Initiative (GEI) Z01 CP 010200, NIH U01 HG004446, and NIH GEI U01 HG 004438.

### SELECT

Research reported in this publication was supported in part by the National Cancer Institute of the National Institutes of Health under Award Numbers U10 CA037429 (CD Blanke), and UM1 CA182883 (CM Tangen/IM Thompson). The content is solely the responsibility of the authors and does not necessarily represent the official views of the National Institutes of Health.

### SMS and REACH

This work was supported by the National Cancer Institute (grant P01 CA074184 to J.D.P. and P.A.N., grants R01 CA097325, R03 CA153323, and K05 CA152715 to P.A.N., and the National Center for Advancing Translational Sciences at the National Institutes of Health (grant KL2 TR000421 to A.N.B.-H.)

### The Swedish Low-risk Colorectal Cancer Study

The study was supported by grants from the Swedish research council; K2015-55X-22674-01-4, K2008-55X-20157-03-3, K2006-72X-20157-01-2 and the Stockholm County Council (ALF project).

### Swedish Mammography Cohort and Cohort of Swedish Men

This work is supported by the Swedish Research Council /Infrastructure grant, the Swedish Cancer Foundation, and the Karolinska Institute’s Distinguished Professor Award to Alicja Wolk.

### UK Biobank

This research has been conducted using the UK Biobank Resource under Application Number 8614 VITAL: National Institutes of Health (K05 CA154337).

### WHI

The WHI program is funded by the National Heart, Lung, and Blood Institute, National Institutes of Health, U.S. Department of Health and Human Services through contracts HHSN268201100046C, HHSN268201100001C, HHSN268201100002C, HHSN268201100003C, HHSN268201100004C, and HHSN271201100004C.

The American Cancer Society funds the creation, maintenance, and updating of the Cancer Prevention Study-II cohort.

PV and LV acknowledge the support by the Grant Agency of the Czech Republic (20-03997S, 21-27902S), by the Grant Agency of the Ministry of Health of the Czech Republic (grants AZV NV18/03/00199 and AZV NV21-03-00506), and Charles University grants Unce/Med/006 and Progress Q28/LF1.

This work was supported by Cancer Research UK [grant number C18281/A29019].

## ASTERISK

We are very grateful to Dr. Bruno Buecher without whom this project would not have existed. We also thank all those who agreed to participate in this study, including the patients and the healthy control persons, as well as all the physicians, technicians and students.

## CCFR

The Colon CFR graciously thanks the generous contributions of their study participants, dedication of study staff, and the financial support from the U.S. National Cancer Institute, without which this important registry would not exist. The authors would like to thank the study participants and staff of the Seattle Colon Cancer Family Registry and the Hormones and Colon Cancer study (CORE Studies).

## CLUE II

We thank the participants of Clue II and appreciate the continued efforts of the staff at the Johns Hopkins George W. Comstock Center for Public Health Research and Prevention in the conduct of the Clue II Cohort Study.

## CORSA

We kindly thank all individuals who agreed to participate in the CORSA study. Furthermore, we thank all cooperating physicians and students and the Biobank Graz of the Medical University of Graz.

## CPS-II

The authors thank the CPS-II participants and Study Management Group for their invaluable contributions to this research. The authors would also like to acknowledge the contribution to this study from central cancer registries supported through the Centers for Disease Control and Prevention National Program of Cancer Registries, and cancer registries supported by the National Cancer Institute Surveillance Epidemiology and End Results program.

## Czech Republic CCS

We are thankful to all clinicians in major hospitals in the Czech Republic, without whom the study would not be practicable. We are also sincerely grateful to all patients participating in this study.

## DACHS

We thank all participants and cooperating clinicians, and everyone who provided excellent technical assistance.

## EPIC

Where authors are identified as personnel of the International Agency for Research on Cancer/World Health Organization, the authors alone are responsible for the views expressed in this article and they do not necessarily represent the decisions, policy or views of the International Agency for Research on Cancer/World Health Organization.

## Harvard cohorts

The study protocol was approved by the institutional review boards of the Brigham and Women’s Hospital and Harvard T.H. Chan School of Public Health, and those of participating registries as required. We acknowledge Channing Division of Network Medicine, Department of Medicine, Brigham and Women’s Hospital as home of the NHS. The authors would like to acknowledge the contribution to this study from central cancer registries supported through the Centers for Disease Control and Prevention’s National Program of Cancer Registries (NPCR) and/or the National Cancer Institute’s Surveillance, Epidemiology, and End Results (SEER) Program. Central registries may also be supported by state agencies, universities, and cancer centers. Participating central cancer registries include the following: Alabama, Alaska, Arizona, Arkansas, California, Colorado, Connecticut, Delaware, Florida, Georgia, Hawaii, Idaho, Indiana, Iowa, Kentucky, Louisiana, Massachusetts, Maine, Maryland, Michigan, Mississippi, Montana, Nebraska, Nevada, New Hampshire, New Jersey, New Mexico, New York, North Carolina, North Dakota, Ohio, Oklahoma, Oregon, Pennsylvania, Puerto Rico, Rhode Island, Seattle SEER Registry, South Carolina, Tennessee, Texas, Utah, Virginia, West Virginia, Wyoming. The authors assume full responsibility for analyses and interpretation of these data.

## Kentucky

We would like to acknowledge the staff at the Kentucky Cancer Registry.

## LCCS

We acknowledge the contributions of Jennifer Barrett, Robin Waxman, Gillian Smith and Emma Northwood in conducting this study.

## NCCCS I & II

We would like to thank the study participants, and the NC Colorectal Cancer Study staff.

## PLCO

The authors thank the PLCO Cancer Screening Trial screening center investigators and the staff from Information Management Services Inc and Westat Inc. Most importantly, we thank the study participants for their contributions that made this study possible. Cancer incidence data have been provided by the District of Columbia Cancer Registry, Georgia Cancer Registry, Hawaii Cancer Registry, Minnesota Cancer Surveillance System, Missouri Cancer Registry, Nevada Central Cancer Registry, Pennsylvania Cancer Registry, Texas Cancer Registry, Virginia Cancer Registry, and Wisconsin Cancer Reporting System. All are supported in part by funds from the Center for Disease Control and Prevention, National Program for Central Registries, local states or by the National Cancer Institute, Surveillance,

Epidemiology, and End Results program. The results reported here and the conclusions derived are the sole responsibility of the authors.

## SEARCH

We thank the SEARCH team

## SELECT

We thank the research and clinical staff at the sites that participated on SELECT study, without whom the trial would not have been successful. We are also grateful to the 35,533 dedicated men who participated in SELECT.

## WHI

The authors thank the WHI investigators and staff for their dedication, and the study participants for making the program possible. A full listing of WHI investigators can be found at: http://www.whi.org/researchers/Documents%20%20Write%20a%20Paper/WHI%20Investigator%20Short%20List.pdf

The authors express sincere appreciation to all Cancer Prevention Study-II participants, and to each member of the study and biospecimen management group. The authors would like to acknowledge the contribution to this study from central cancer registries supported through the Centers for Disease Control and Prevention’s National Program of Cancer Registries and cancer registries supported by the National Cancer Institute’s Surveillance Epidemiology and End Results Program.

