## Supplementary methods and figures for "Probing the diabetes and colorectal cancer relationship using gene – environment interaction analyses"

**Two-step method**

We implemented a hybrid two-step method that prioritizes potential interaction loci by weighting GxE tests (step 2) based on the ranks of the EDGE statistic (step 1). Our approach uses a weighted hypothesis testing framework, which uses step 1 ranks to prioritize and partition variants into exponentially larger bins of fixed sizes and increasingly more stringent step 2 significance thresholds (1). As originally proposed (1), a limitation of the standard weighted testing is that the top bins are often filled with correlated markers from the same loci. To address this issue, we developed a modified weighted testing method that accommodates bins of varying sizes while accounting for linkage disequilibrium and properly controlling the type I error (2). Specifically, variants are partitioned into bins based on step 1 p-value thresholds, with Bin 1 including SNPS with Step-1 p<5/1M, Bin 2 SNPs with 5/1M < p < 15/1M, Bin 3 SNPs with 15/1M < p < 35/1M, etc. For step 2, significance of the standard 1-d.f. GxE test statistic was evaluated at a bin-specific significance threshold that preserves the overall familywise error rate while accounting for the influx of correlated markers into each bin.

**Prediction of regulatory impact of candidate non-coding variants**

For each biosample, the SVMs were trained on 120,000 genomic regions. Positively labeled regions were 1 kb DNA sequences centered on the summits of the 60,000 most significant DNase-seq peaks from MACS2+IDR, and 60,000 1 kb negatively labeled sequences were randomly sampled from the GRCh38 reference genome such that they did not overlap IDR and naive overlap DNase-seq peaks and were matched for GC content to the positive regions.

The resulting trained models for each of the five DNASE-seq datasets were used to score all variants on the Haplotype Reference Consortium (HRC) imputed panel (n=39,117,106). For each variant along the HRC panel, we centered a 1 kb sequence interval and obtained SVM model predictions for the reference and alternate alleles. The difference in model predictions of accessibility (prediction for alternate allele - prediction for reference allele) are the in-silico mutagenesis scores (ISM), or variant effect scores. We confirmed that the ISM scores for the HRC panel were normally distributed using the Kolmogorov-Smirnov and Shapiro Wilkes tests (p-values>0.10) and derived Z -scores. Variants with ISM scores greater than 1.65 or less than -1.65, representing a 90% confidence interval, were determined to have significant effects. A single score was obtained for each HRC variant by taking the maximum of the absolute values of the GKMexplain delta scores across the five models.

The lead GWAS variants were LD-expanded (500 kb window, *R*^2^ thresholded at 0.20) using PLINK (1.9) (3) based on the 1000 genomes phase 3 fileset from the cog-genomics site (<https://www.cog-genomics.org/plink/2.0/resources#1kg_phase3>) (4), which was filtered to separate individuals of CEU ancestry. Using the SVM models trained on each of the five DNase-seq datasets, we scored the LD-expanded significant locus and predicted ISM effects on chromatin accessibility. We further inferred the contribution scores of each nucleotide in the input sequences to the output prediction of the SVM models using the GKMexplain algorithm (5). For each sequence containing a candidate variant, we computed GkmExplain scores for the sequence containing the reference allele and the sequence containing the alternate allele. For each candidate variant, a deltaGKMexplain score was computed by subtracting the GKMexplain score for the 1 kb vector of GKMexplain scores of the sequence with the reference allele from the 1 kb vector of GKMexplain scores of the sequence with the alternate allele. The TomTom algorithm (6) was used to identify likely motif matches for subsequences with high deltaGKMexplain scores. The support vector machine LS-GKM + GKMexplain workflow source code is available on github: https://github.com/kundajelab/SVM_pipelines.

Additionally, to assess tissue-specificity we intersected a compendium of ten groups of tissue-specific regulatory annotations for histone modifications (H3K4me1, H3K4me3, H3K9ac, and H3K27ac) obtained from various resources (7, 8), and previously derived by Finucane et al. and assessed whether our significant findings coincide with open chromatin regions (9).

**Determination of gene-enhancer links**

Rare variants were constructed into sets based on genetic region and enhancer sites. The latter were identified from several annotation sources. Gene-enhancer links within the GRCh37 genome build were identified as described below. GWAS variants found within the resulting gene loci or enhancer regions were selected for downstream analysis. Gencode (v31) GRCh37 annotations were downloaded from EBI (ftp://ftp.ebi.ac.uk/pub/databases/gencode/Gencode_human/release_31/GRCh37_mapping/gencode.v31lift37.annotation.gtf.gz). The transcription start sites (TSS) regions were identified and regions within +/- 5kb of the TSS were obtained. H3K27ac histone ChIP-seq datasets in COLO205, HCT116, and SW480 cell lines, healthy colon and tumor primary samples were generated by Scacheri et al (10). The MACS2 peaks within these datasets were intersected with TSS+/-100 bp regions to identify active genes within each of the five datasets. Peaks within 5kb of active TSS sites were then identified, indicating active enhancers within the vicinity of active genes.

Reg2Map Honeybadger2 stringent (P-value: <1x10^-10^) promoter and enhancer annotations at (https://personal.broadinstitute.org/meuleman/reg2map/HoneyBadger2_release/DNase/p10/enh/BED_files_per_sample/) were obtained for digestive ("E075", "E077", "E079", "E084", "E085", "E092", "E094", "E101", "E102", "E106", "E109", "E110") and immune ( "E030", "E031", "E032", "E034", "E044") cell types. These annotations were overlapped with TSS+/-5KB regions for active TSS sites, providing annotations of TSS-specific enhancers for each of the above-mentioned cell types.

Additionally, the gene-enhancer annotations were augmented with the Union Links file (wget https://personal.broadinstitute.org/anshul/projects/roadmap/integrative/enh_gene_links/Union_link_sig_FDR05_Sep.txt.gz) from Roadmap.

**Supplementary figures**

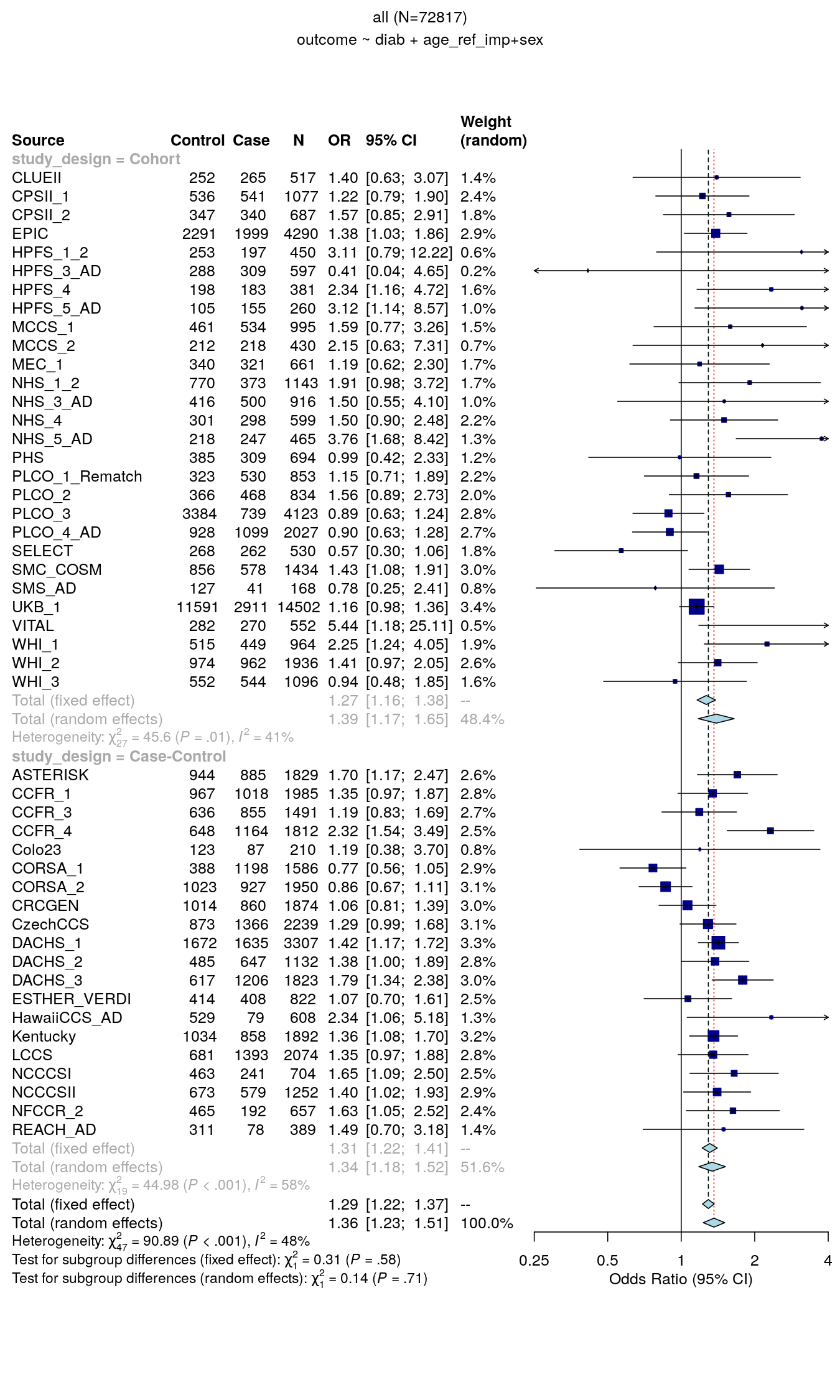

**Supplementary Figure S1**. Forest plot of the association between diabetes and colorectal cancer risk for all studies included in the gene-diabetes interaction analysis for colorectal cancer risk adjusted for age at baseline and sex.

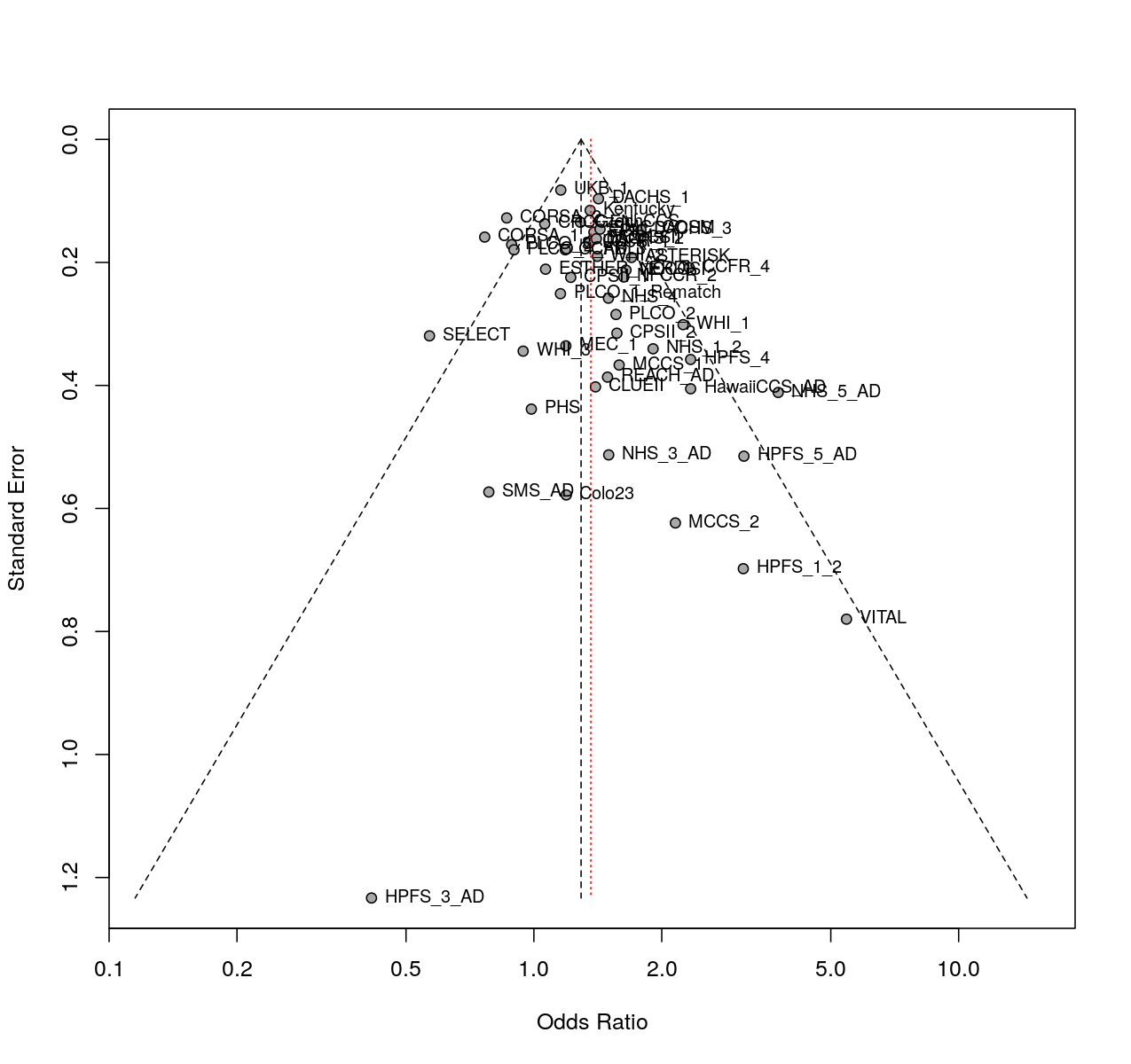

**Supplementary Figure S2**. Funnel plot of the studies included in the gene-diabetes interaction analysis for colorectal cancer risk adjusted for age at baseline and sex.

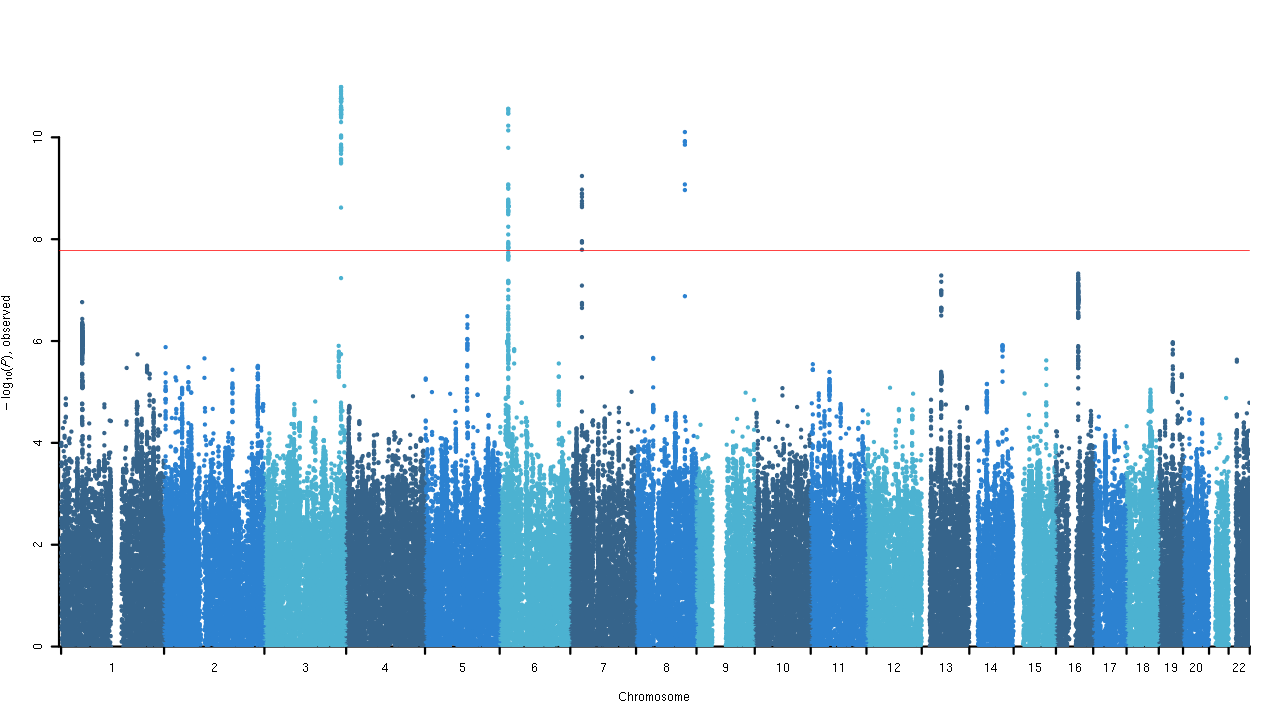

A)

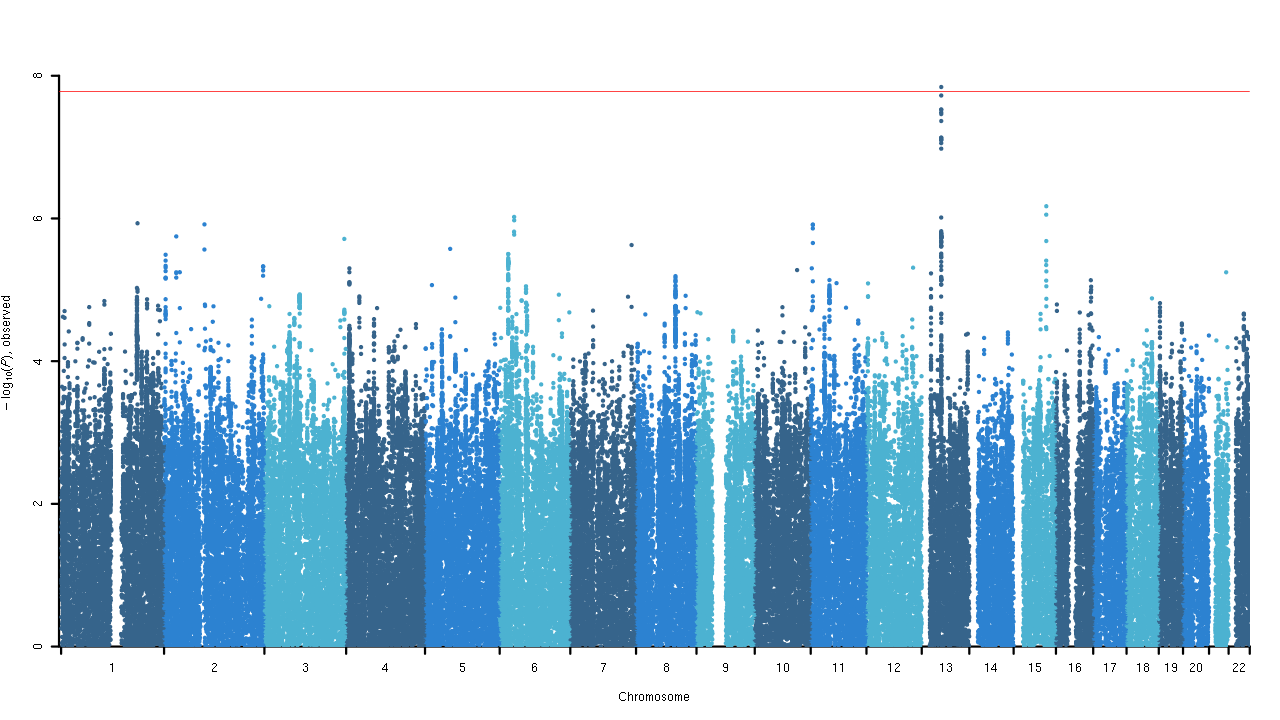

B)

**Supplementary Figure S3**. Manhattan plot of the gene-environment interaction of (A) 3-degrees of freedom joint test for the interaction between genetic variants and diabetes on colorectal cancer risk and the association between genetic variants and colorectal cancer and the association between genetic variants and diabetes risk and (B) 2-degrees of freedom joint test for the interaction between genetic variants and diabetes on colorectal cancer risk and the association between genetic variants and colorectal cancer risk adjusted for age at baseline, sex, study, genotyping platform, and the first three principal components. Known genome-wide significant loci for colorectal cancer have been removed from these analyses.

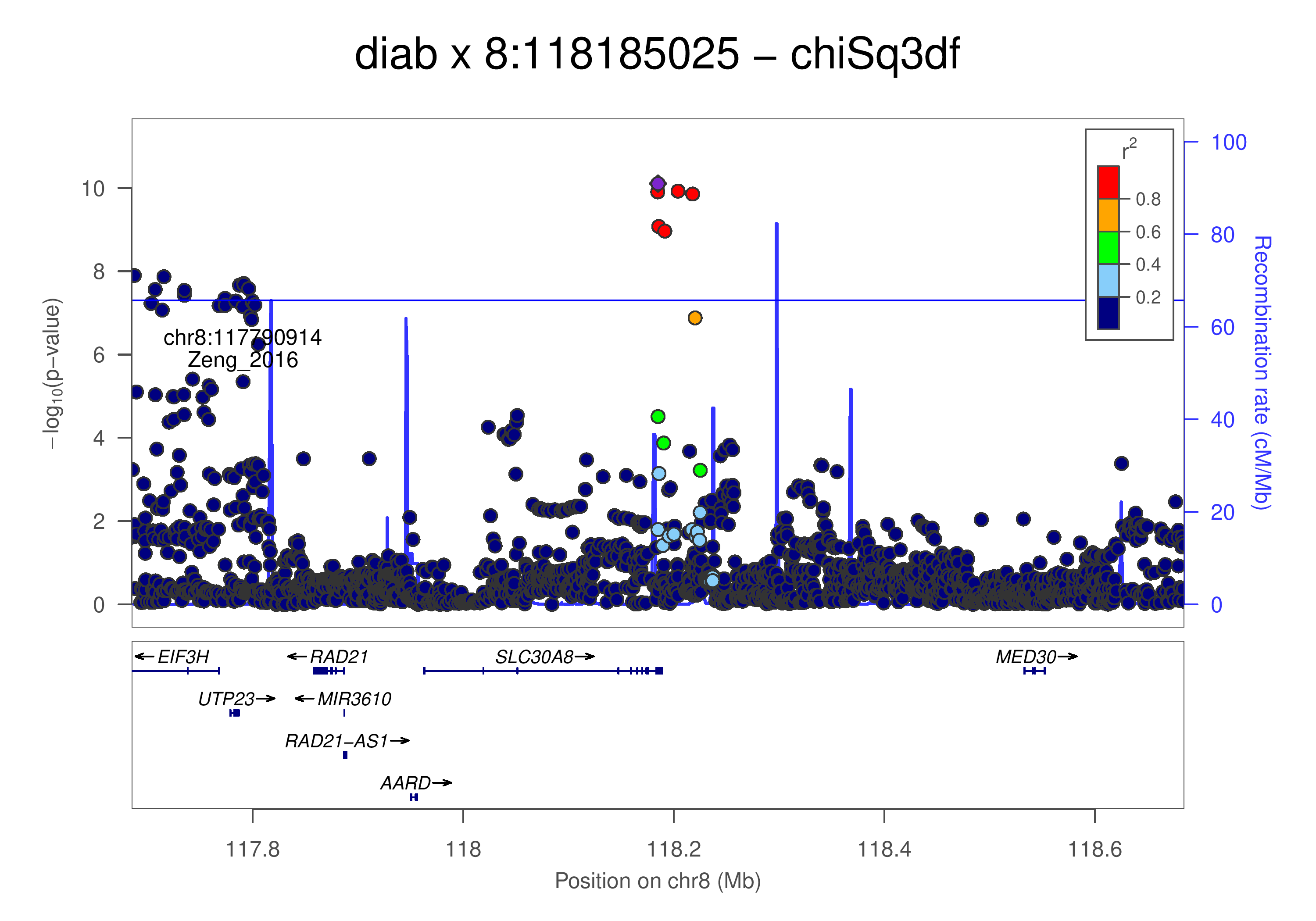

A)

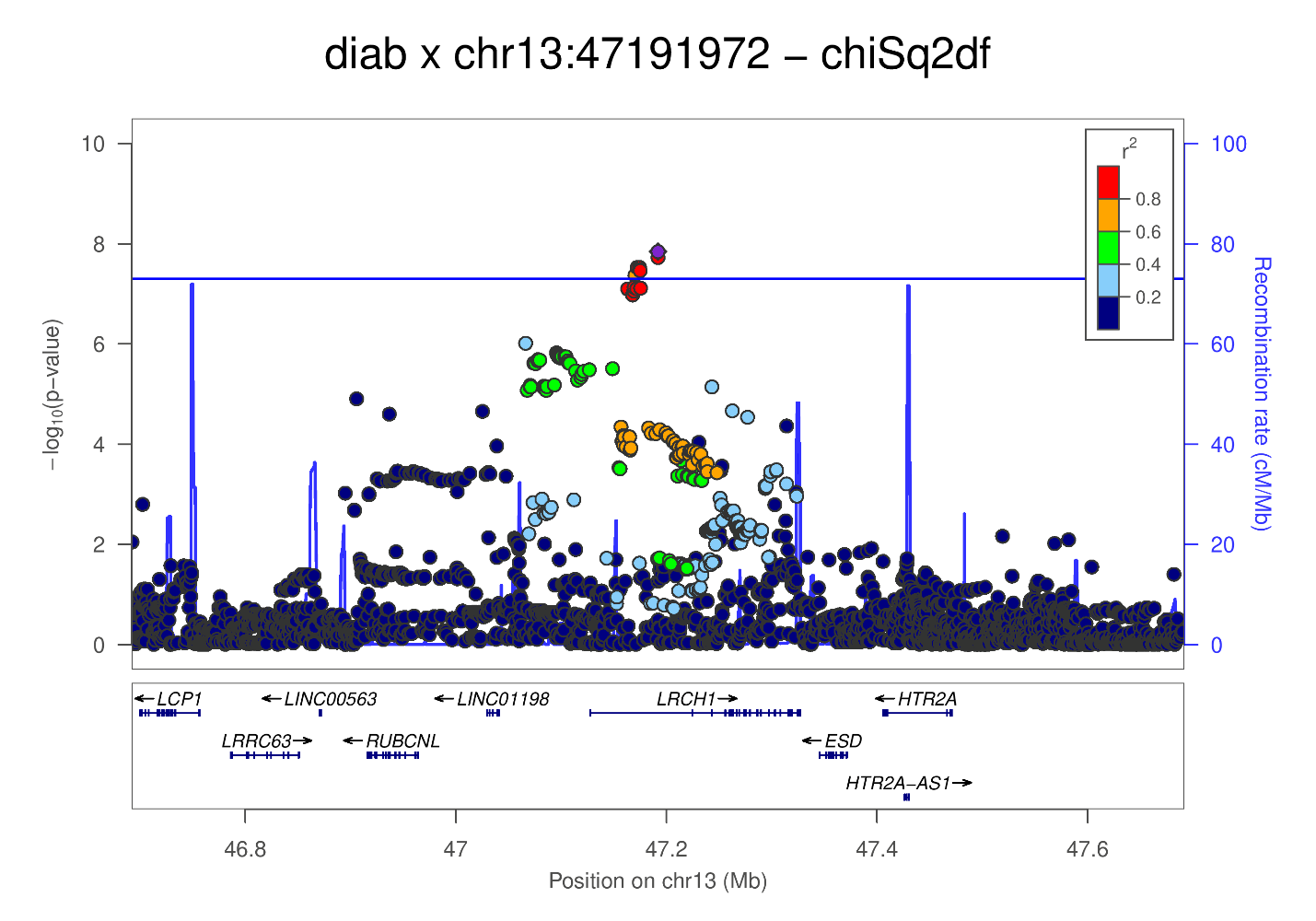

B)

**Supplementary Figure S4**. Regional plots for the significant findings (A) rs3802177, and B) rs9526201) of the gene-diabetes interaction analysis for colorectal cancer risk.

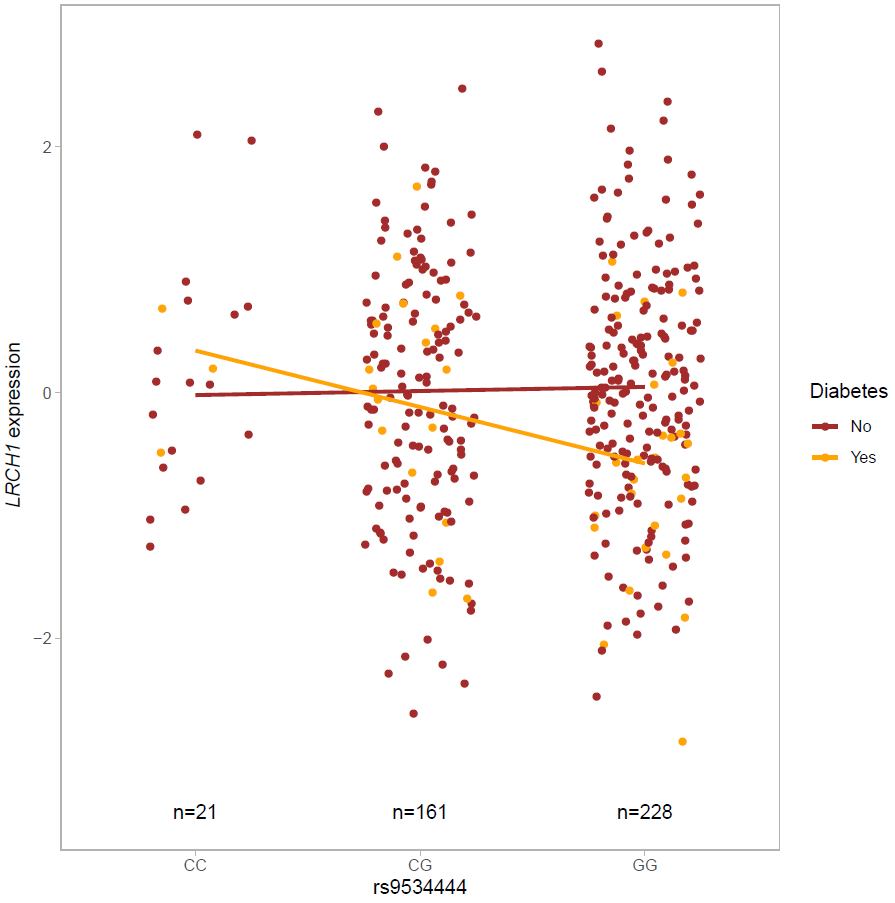

**Supplementary Figure S5**. Interaction plot of variant rs9534444 (linkage disequilibrium *R*^2^: 0.52 with rs9526201) with diabetes in relation to *LRCH1* gene expression in BarcUVa-Seq samples (N: 49 individuals with diabetes; N: 361 without diabetes).

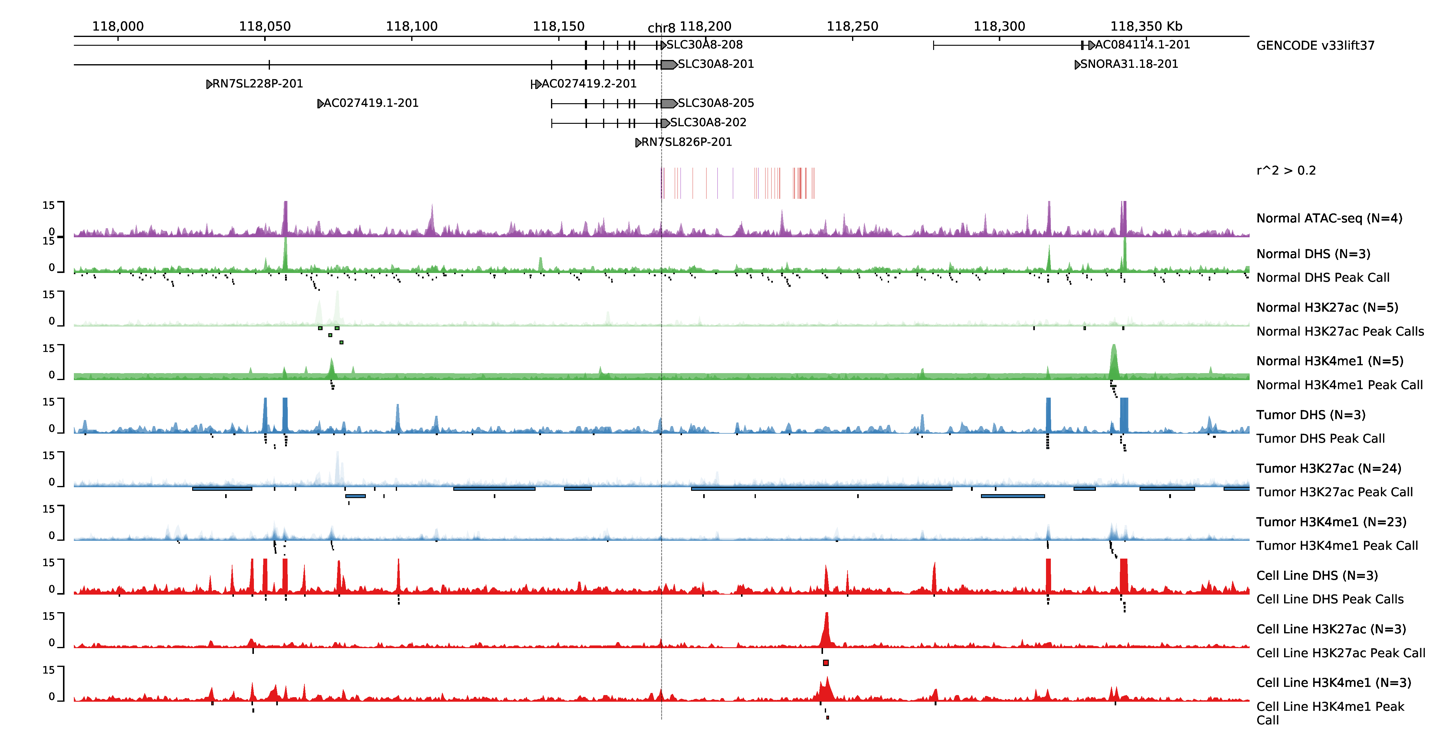

A)

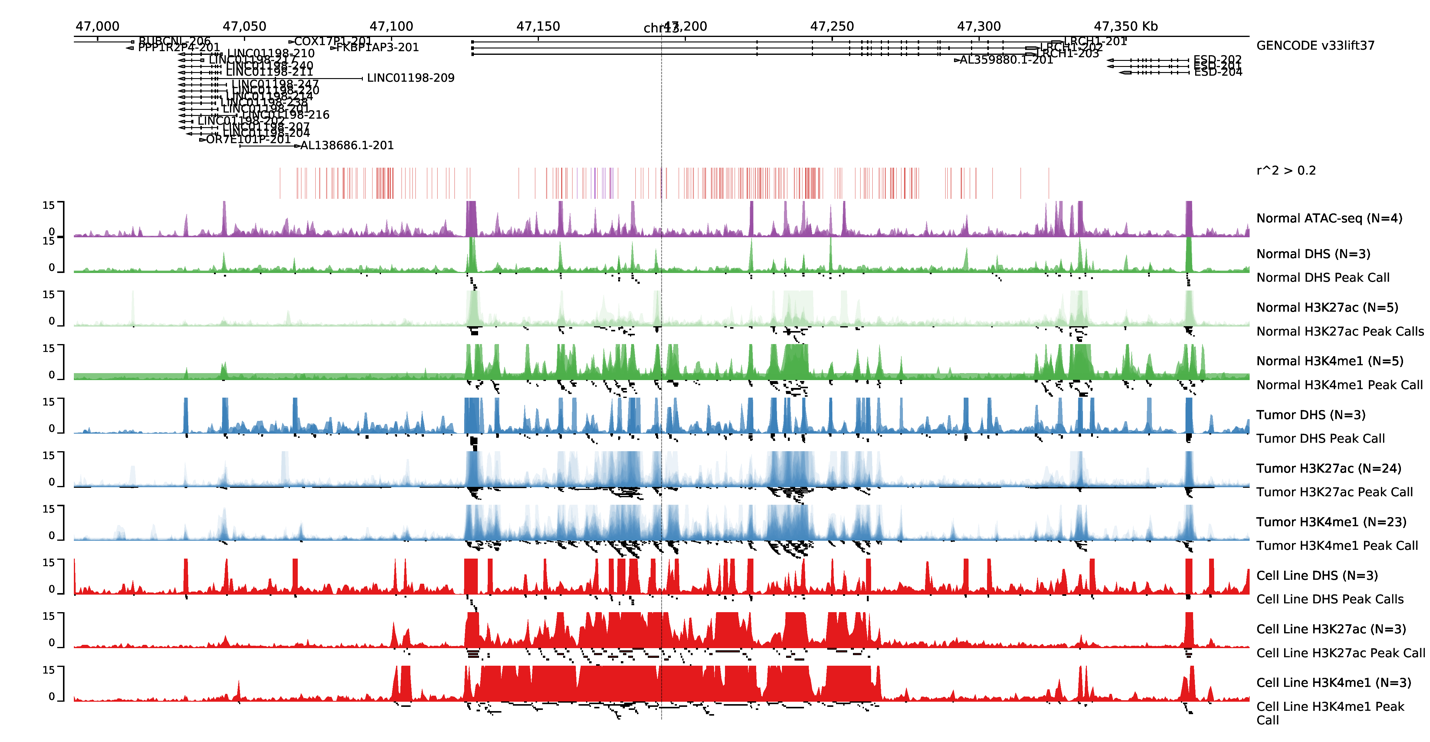

B)

**Supplementary Figure S6**. Chromatin accessibility assays highlighting peaks for variants with *R*^2^ > 0.2 within 500kb of the significant findings (A) rs3802177, and B) rs9526201) in the gene-diabetes interaction analysis for colorectal cancer risk. Top panel indicates GENCODE reference genes (GRCh37).

| A) | |
| --- | --- |
| 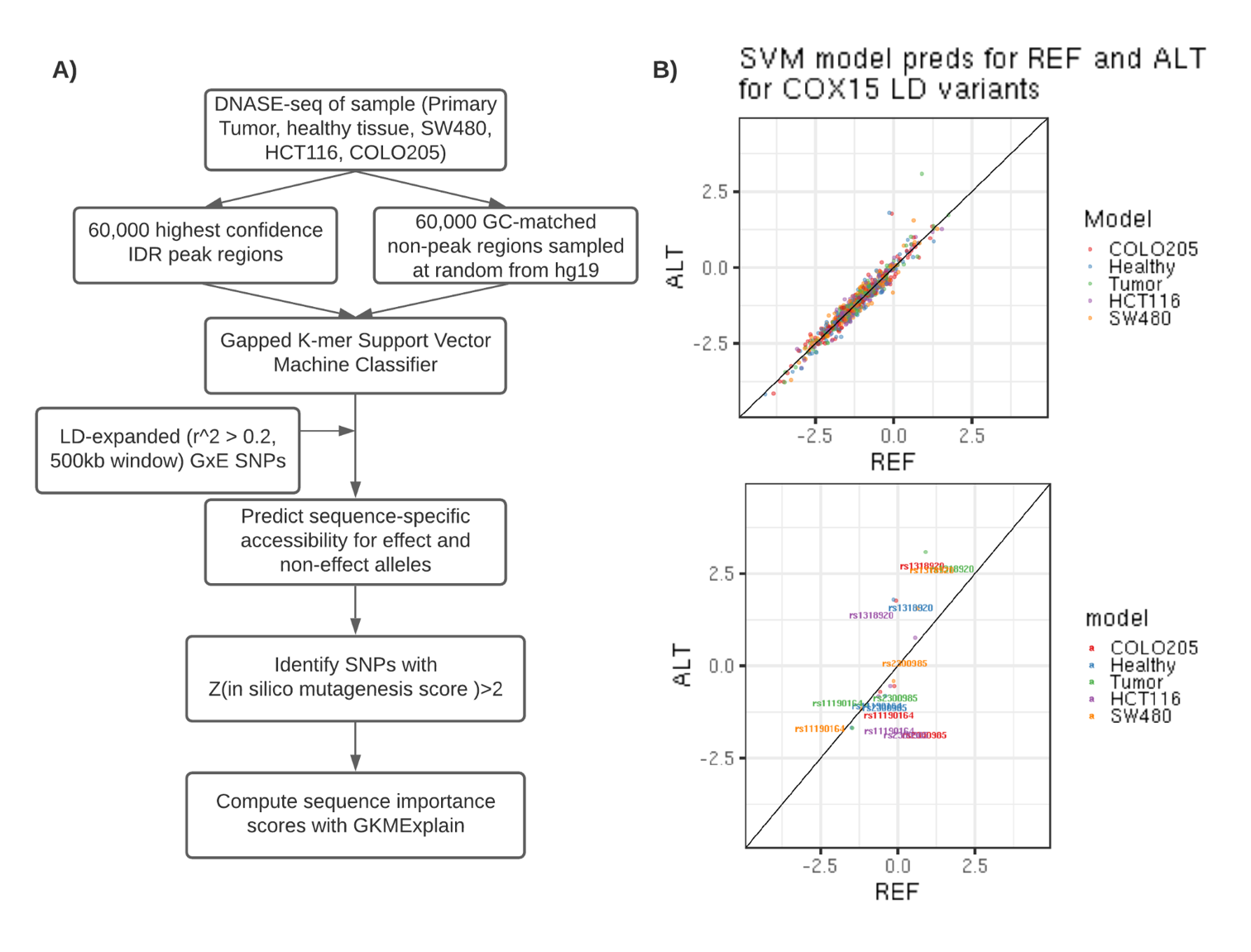 | |
| B) | C) |
| 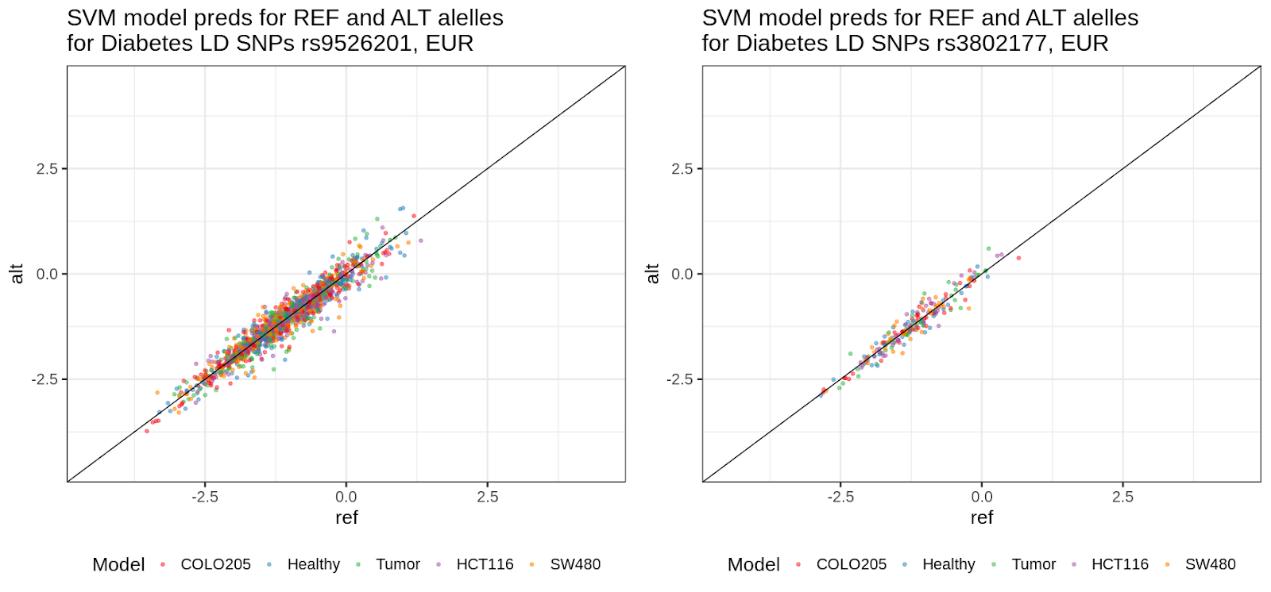 | 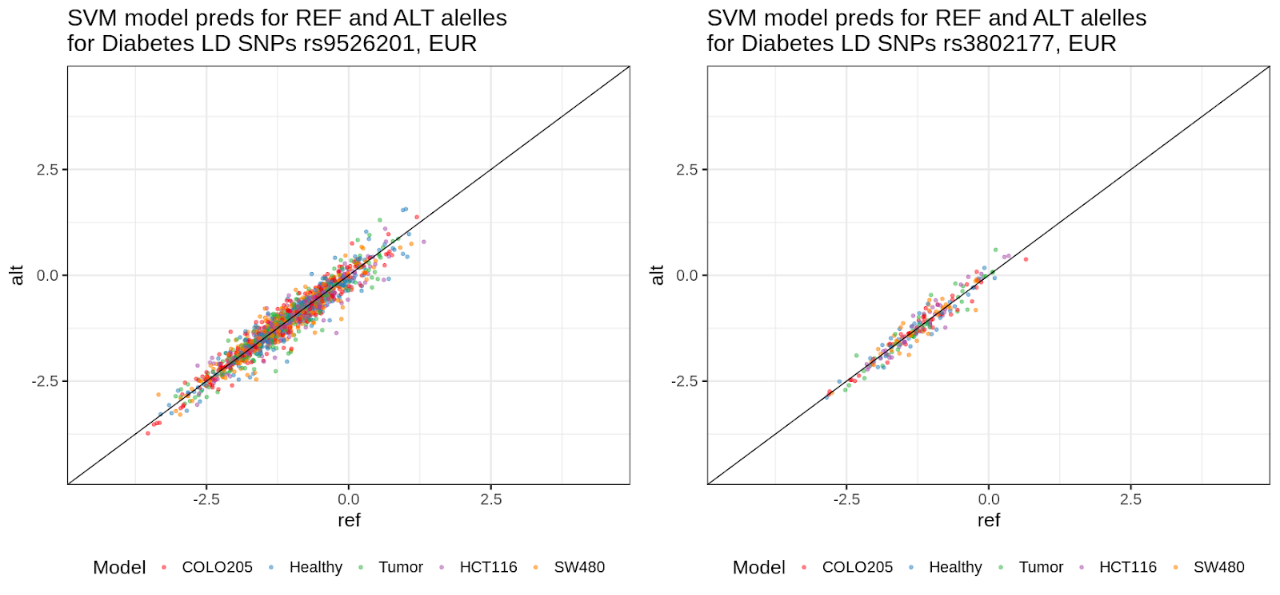 |
| 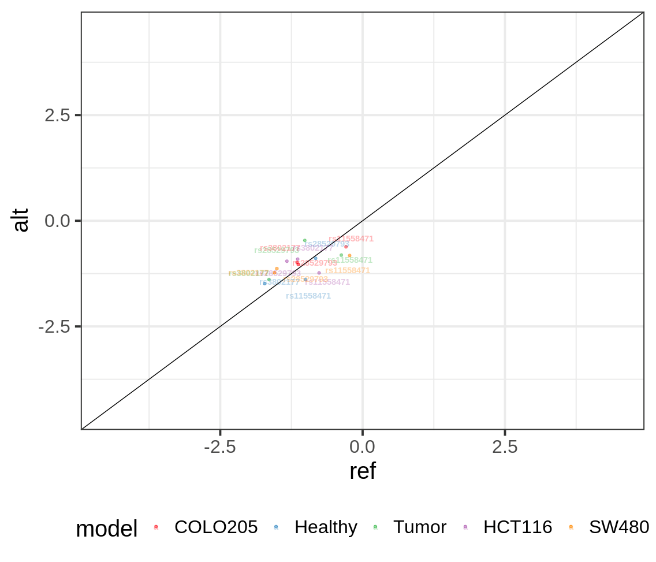 | 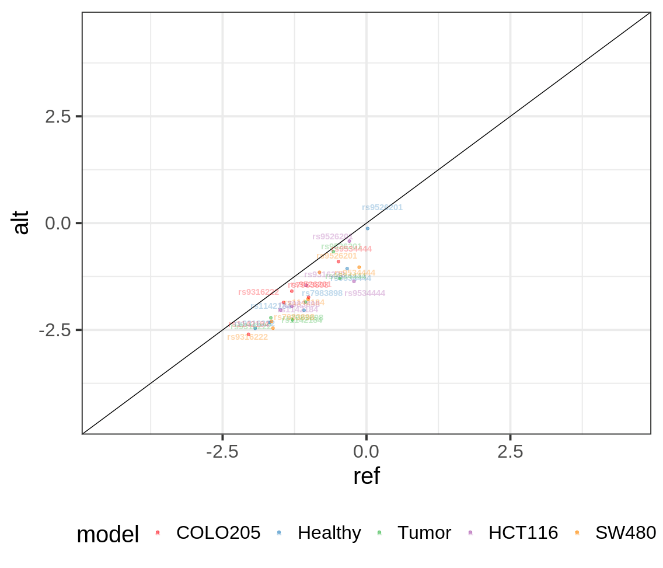 |

**Supplementary Figure S7.** Support vector machine learning model pipeline to predict functional effects of linked variants within the rs3802177 and rs9526201 region. **A)** Analysis pipeline for SVM classifier development and linked variant scoring. **B)** SVM test set predictions for reference and alternate alleles for 41 variants with *R*^2^ > 0.2 within 500kb of the *SLC30A8* rs3802177. Bottom panel highlights reference and alternate predictions for rs3802177. **C)** SVM test set predictions for reference and alternate alleles for 321 variants with *R*^2^ > 0.2 within 500kb of the *LRCH1* rs9526201 variant. Bottom panel highlights reference and alternate predictions for rs9526201.

| A) | B) | C) |
| --- | --- | --- |
| 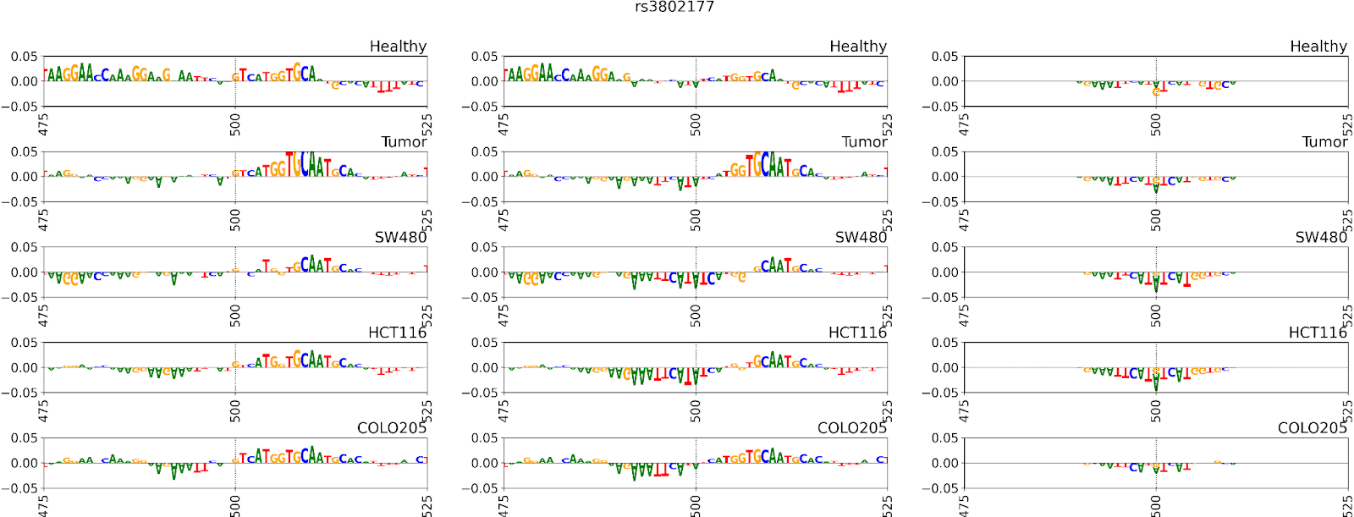 | | |
| D) | E) | F) |
| 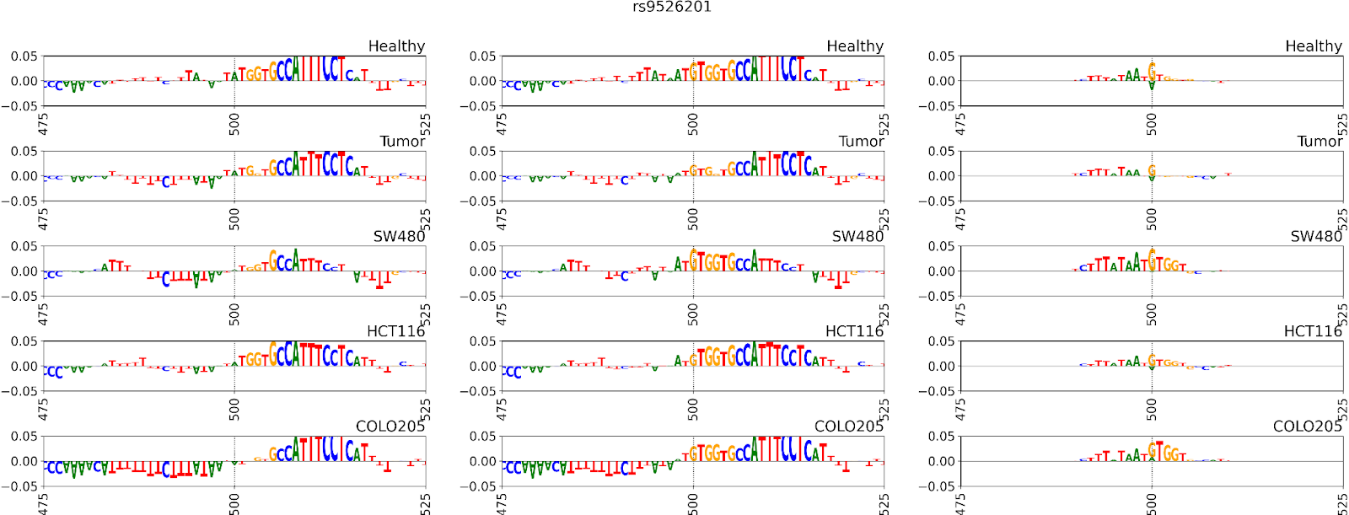 | | |
| G) | H) | I) |
| 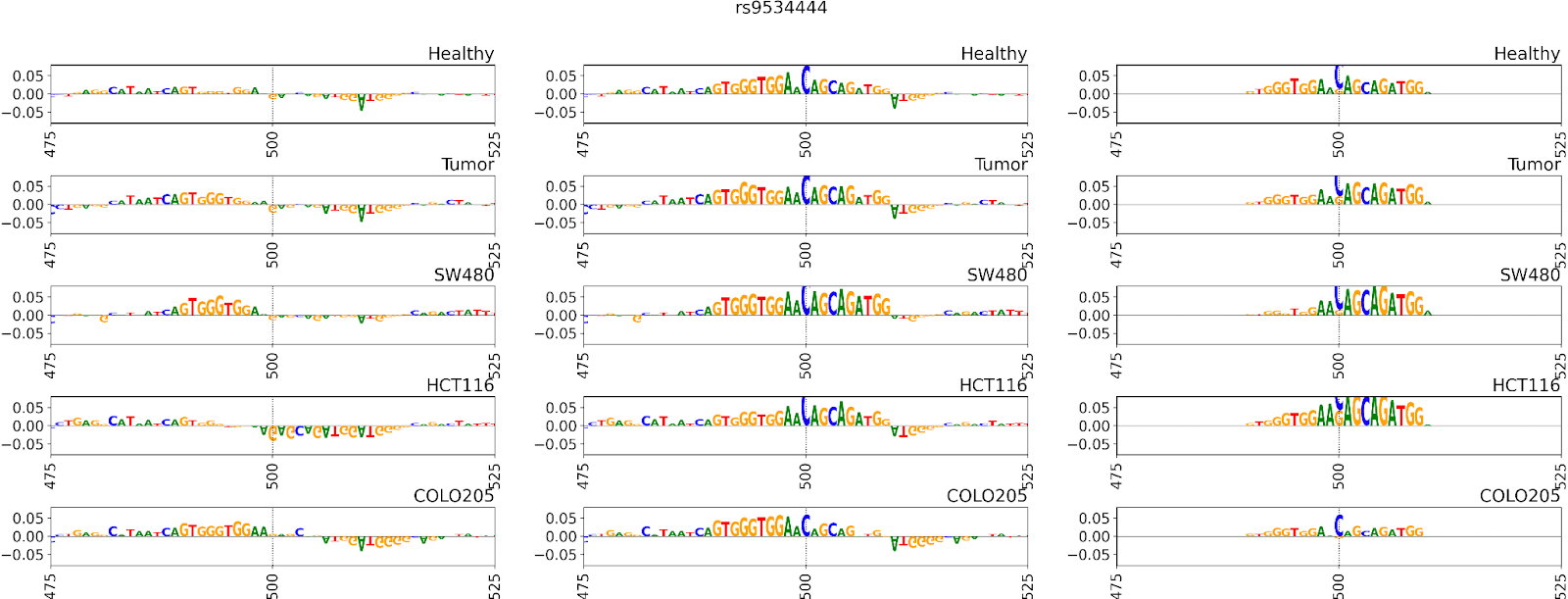 | | |
| **Supplementary Figure S8.** GkmExplain sequence importance scores within +/- 50 bp of the variants of interest *SLC30A8* and *LRCH1* region. Scores are derived from SVM models in healthy and tumor primary tissue samples as well as SVM models in cell lines SW480, HCT116, COLO205. **A-C)** Importance scores for rs3802177, with A) indicating allele G (alternate), B) indicating allele A (reference), and C) indicating the delta score for the A allele minus the G allele. **D-F)** Importance scores for rs9526201, with D) indicating allele A (alternate), E) indicating allele G (reference), and F) indicating the delta score for the G signal minus the A signal. **G-I)** Importance scores for rs9534444, with G) indicating allele G (alternate), H) indicating the reference allele C (reference), and I) indicating the delta score for C signal minus the G signal. | | |
